# Acute respiratory infections due to antibiotic-nonsusceptible *Streptococcus pneumoniae* in United States adults

**DOI:** 10.1101/2025.04.04.25325277

**Authors:** Laura M King, Kristin L Andrejko, Miwako Kobayashi, Wei Xing, Adam L Cohen, Joseph A Lewnard

## Abstract

**Background:** We aimed to estimate the burden of antibiotic-nonsusceptible non-bacteremic pneumonia and sinusitis due to *Streptococcus pneumoniae* (pneumococcus) in US adults (≥18 years).

**Methods:** We estimated antibiotic-nonsusceptible pneumococcal sinusitis and non-bacteremic pneumonia incidence as products of non-bacteremic pneumococcal pneumonia and sinusitis incidence rates, serotype distribution, and serotype-specific antimicrobial nonsusceptibility prevalences by antibiotic class and guideline-recommended agents from 2016-2019. We derived pneumonia and sinusitis incidence rates from national healthcare utilization surveys and administrative datasets; pneumococcal-attributable attributable percents and serotype distributions from published data; and serotype-specific nonsusceptibility estimates from Active Bacterial Core surveillance data. We evaluated nonsusceptibility for all serotypes and those targeted by 15-, 20- and 21-valent pneumococcal conjugate vaccines (PCV15/20/21).

**Results:** An estimated 16.4% (95% confidence interval 12.8-21.4%) of non-bacteremic pneumococcal pneumonia and 19.0% (14.8-24.9%) of sinusitis cases were nonsusceptible to ≥3 antibiotic classes, translating to 243,521 (179,673-333,675) and 1,844,726 (1,070,763-2,904,089) outpatient visits for pneumonia and sinusitis, respectively, and 10,155 (7,542-13,803) pneumonia hospitalizations annually. An estimated 31.2% (26.6-36.3%) of non-bacteremic pneumococcal pneumonia and 10.5% (9.4-12.0%) of pneumococcal sinusitis cases were nonsusceptible to ≥1 outpatient first-line antibiotic agent. Cases attributable to serotypes targeted by PCV15, PCV20, and PCV21 that were nonsusceptible to ≥3 antibiotic classes accounted for 7.4% (4.7-11.1%), 8.5% (5.8-12.1%), and 12.6% (9.2-17.5%) of all non-bacteremic pneumococcal pneumonia cases, and 8.4% (5.3-12.5%), 9.4% (6.2-13.4), and 14.4% (10.4-20.0%) of all pneumococcal sinusitis cases.

**Conclusions:** We demonstrated high proportions of antibiotic nonsusceptibility in non-bacteremic pneumococcal pneumonia and sinusitis in US adults. PCVs and antibiotic stewardship may mitigate antibiotic nonsusceptibility in pneumococcal disease.

## INTRODUCTION

Antibiotic-nonsusceptible *Streptococcus pneumoniae* (pneumococcus) infections are recognized by the US Centers for Disease Control and Prevention and the World Health Organization as important threats to human health.^1,2^ Acute respiratory infections (ARIs) account for the majority of pneumococcal disease cases among adults^3^ (non-bacteremic pneumonia, sinusitis) and children^4^ (non-bacteremic pneumonia, acute otitis media [AOM]). Additionally, invasive pneumococcal disease (IPD; bacteremic pneumonia, sepsis, meningitis) accounts for substantial morbidity and mortality. In both IPD and ARIs, antibiotic nonsusceptibility may enhance the complexity of patients’ care needs. Meta-analyses of studies of IPD^5^ and pneumococcal pneumonia^6^ found that penicillin-nonsusceptibility was associated with 1.3-fold higher odds of 30-day mortality. However, nonsusceptibility to macrolides (infrequently used in inpatient pneumonia) was not associated with in-hospital mortality in a study of hospitalized Spanish adults.^7^ A US health economic study estimated that nonsusceptibility to penicillin, erythromycin, and fluoroquinolones in pneumococcal pneumonia resulted in an additional 32,000 outpatient visits and 19,000 hospitalizations, resulting in $233 million in excess costs in 2012.^8^ Due to its potential importance in ARI management, nonsusceptibility in pneumococcus is regularly considered in treatment guidelines for syndromes such as pneumonia^9^ and sinusitis.^10,11^

There are >100 recognized pneumococcal serotypes,^12^ with a small proportion of these accounting for most disease. Antibiotic-nonsusceptibility varies widely across serotypes.^13–15^ For instance, among IPD isolates from US Active Bacterial Core surveillance (ABCs) >50% of isolates from serotypes 19A, 33F, 35B, and 15A were nonsusceptible to ≥1 antibiotic class, while nonsusceptibility was rare among serotypes 8, 20, and 38.^13^ Although studies of nonsusceptibility in pneumococcal ARIs are limited, serotypes generally show similar patterns of antibiotic susceptibility in studies of invasive and non-invasive isolates, with some variation by class.^15,16^ Historically, 7-, 10-, and 13-valent pneumococcal conjugate vaccines (PCVs; PCV7/10/13), targeting serotypes prevalent in IPD and those nonsusceptible to antibiotics, were associated with decreases in antibiotic-nonsusceptible pneumococcal infections^13,17,18^ and colonization.^19^

Currently, newly-licensed pneumococcal conjugate vaccines (PCVs) targeting 15, 20, and 21 serotypes (PCV15/20/21) are recommended for US adults meeting eligibility criteria.^20^ Antibiotic nonsusceptibility patterns and the contributions of distinct serotypes to pneumococcal disease burden are important considerations for clinical guidelines and next-generation vaccine formulations. While nonsusceptibility in IPD is well-characterized, the significant burden of pneumococcal ARIs underscores the importance of better understanding antibiotic-nonsusceptibility in these infections. We aimed to estimate the burden of non-bacteremic pneumonia and sinusitis associated with antibiotic-nonsusceptible *S. pneumoniae* in US adults.

## METHODS

### Overview

We estimated the incidence of antibiotic nonsusceptible (intermediate susceptibility or resistant) pneumococcal sinusitis and non-bacteremic pneumonia among adults using the following inputs: incidence rates for all-cause non-bacteremic pneumonia and sinusitis; the proportion of cases for each of these syndromes which were expected to be attributable to pneumococcus; the distribution of serotypes associated with non-bacteremic pneumococcal pneumonia and sinusitis among adults; and the prevalence of antimicrobial nonsusceptibility within each pneumococcal serotype over the years 2016-2019. We selected 2016-2019 to ensure multiple, continuous years of data without perturbations from COVID-19 pandemic-associated shifts in healthcare seeking and pneumococcal disease.^21^ We estimated burdens for all adults and within age-group strata (18-49, 50-64 and ≥65 years) for each syndrome. We derived incidence rates of non-bacteremic pneumonia (for cases managed across inpatient and outpatient settings) and sinusitis (cases managed in outpatient settings only) from national healthcare utilization surveys and administrative datasets. We estimated attributable fractions of cases due to pneumococcus and pneumococcal serotype distributions using published data. Lastly, we obtained serotype-specific antibiotic nonsusceptibility estimates from active population and laboratory based surveillance. We evaluated nonsusceptibility for all serotypes, serotypes targeted by PCVs (PCV15, PCV20, and PCV21), and non-vaccine type serotypes (NVTs; i.e., those not included in either PCV20 nor PCV21).

This activity was reviewed by CDC, deemed research not involving human subjects, and was conducted consistent with applicable federal law and CDC policy. See e.g., 45 C.F.R. part 46, 21 C.F.R. part 56; 42 U.S.C. §241(d); 5 U.S.C. §552a; 44 U.S.C. §3501 et seq.

### Incidence of pneumococcal ARIs among adults

We estimated annual counts of all-cause pneumonia hospitalizations from the 2019 National Inpatient Sample using *International Classification of Disease, 10*^*th*^ *revision, Clinical Modification* (ICD-10-CM) codes (**Table S1**).^22^ We multiplied the resulting estimate by the proportion of hospitalized pneumonia cases attributable to pneumococcus, as derived in a study of adults hospitalized with pneumonia in two large regional hospital systems in the Southeastern United States (Pneumococcal pNeumonia Epidemology, Urine serotyping, and Mental Outcomes [PNEUMO];^3^ **Table S2**). From this estimate, we subtracted the number of bacteremic pneumococcal pneumonia cases among adults overall and by age group based on 2019 ABCs data^23^, thus obtaining the total number of hospitalized non-bacteremic pneumococcal pneumonia cases. We divided case counts by corresponding 2019 population bridged-race census estimates^24^ to generate incidence rates.

We estimated incidence rates of outpatient visits for all-cause pneumonia and sinusitis using data from the 2016 and 2019 National Ambulatory and National Hospital Ambulatory Medical Care Surveys (NAMCS/NHAMCS) and 2016-2019 Meritage MarketScan Commercial and Medicaid datasets (MarketScan). Following methods from previous studies,^4,25–27^ we derived incidence rates for outpatient physician office and emergency department visits using NAMCS/NHAMCS, and visit incidence rates for all other outpatient settings from MarketScan. Total incidence rates were calculated as the sum of rate estimates for physician office and emergency department visits (from NAMCS/NHAMCS) and visits for all other settings (from MarketScan data). Overall MarketScan incidence rates were derived as the weighted average of estimates from the Commercial and Medicaid datasets, with weights based on national estimates of private insurance versus other coverage among adults.^28^ Incidence rates were estimated by age group: 18-49, 50-64, and ≥65 years. We multiplied incidence rate estimates by previously-published estimates of the proportions of sinusitis^4^ and pneumonia^3^ cases attributable to pneumococcus. We propagated uncertainty by fitting age- and condition-specific burden estimates to Gamma distributions. We multiplied incidence rates by 2019 bridged-race census estimates^24^ to obtain national counts.

### Pneumococcal serotype distribution

We estimated serotype distributions in adult non-bacteremic pneumococcal pneumonia and sinusitis using methods consistent with previous studies.^27^ For both syndromes, we used data on serotype-specific frequencies from the PNEUMO study,^3^ which used a serotype-specific urinary antigen detection (SSUAD) assay to identify 30 pneumococcal serotypes. For additional serotypes not captured via the PNEUMO SSUAD, we extrapolated serotype-specific frequencies (based on a denominator of all cases involving non-SSUAD serotypes) from proportions observed in 2015-2019 ABCs data. We used Markov chain Monte Carlo to sample from the distribution of serotype-specific proportions, defined as a Dirichlet distribution within each age group (18-49, 50-64, ≥65 years).^27^ We assumed that serotype distributions were consistent for outpatient- and inpatient-managed non-bacteremic pneumonia.

As little data on serotype distribution in pneumococcal sinusitis is available, we additionally conducted a sensitivity analysis applying serotype distribution in pediatric ARIs (AOM and pneumonia) to adult sinusitis. Pediatric ARI serotype distribution was based on a meta-analysis of studies of serotype distribution in pediatric ARIs in high-income countries after PCV13 implementation.^27^

### Serotype-specific antibiotic nonsusceptibility

We estimated the proportion of antibiotic-nonsusceptible cases, by serotype, using ABCs data from 2016-2019. Although limited to IPD, ABCs data provided comprehensive estimates across a large number of serotypes and age groups not available via other data sources.^29^ We defined nonsusceptible isolates as those classified as intermediate susceptibility or resistant based on Clinical and Laboratory Standards Institute (CLSI) breakpoints.^30^ ABCs nonsusceptibility data were stratified by age groups 18-64 and ≥65 years.

We categorized nonsusceptibility by drug class (cephalosporins, lincosamides, antifolates, fluoroquinolones, linezolids, carbapenems, penicillins, tetracyclines, glycopeptides, macrolides) based on nonsusceptibility to specific agents within each class (**Table S3**). To evaluate the potential for treatment failure, we also categorized antibiotic-nonsusceptibility based on nonsusceptibility to guideline-recommended antibiotic agents or classes for the management of pneumonia and sinusitis in adults, to >1 antibiotic class, and to ≥3 antibiotic classes (i.e., multidrug-resistant). For pneumonia, we evaluated nonsusceptibility to guideline-recommended agents as nonsusceptibility to ≥1 outpatient first-line antibiotic agent, ≥2 first-line outpatient antibiotic agents, and ≥1 inpatient first-line antibiotic agent. We defined pneumonia outpatient first-line antibiotics as amoxicillin, doxycycline, and erythromycin (as a proxy for all macrolides), which are agents recommended for the outpatient management of pneumonia in patients without comorbidities or risk factors for complicated infections.^9^ We defined pneumonia first-line inpatient antibiotics as agents recommended for inpatient management of pneumonia or outpatient management of complex cases: cephalosporins (cefotaxime, ceftriaxone, and cefuroxime) and levofloxacin (as a proxy for all respiratory fluoroquinolones).^9^ For sinusitis, we evaluated nonsusceptibility to the recommended first-line antibiotic agent (amoxicillin) and to recommended alternative agents (doxycycline, levofloxacin, clindamycin, and cephalosporins).^10,11^ Although amoxicillin-clavulanate rather than amoxicillin is recommended as a first-line treatment for pneumonia and sinusitis, ABCs nonsusceptibility testing is limited to amoxicillin. We propagated uncertainty by defining the prevalence of nonsusceptibility within each age group and serotype stratum as a Beta distribution, parameterized by counts of nonsusceptible and susceptible isolates.

Where serotype-specific nonsusceptibility proportions were available in only one age group, we applied that proportion across all ages. For serotypes without nonsusceptibility values (i.e., serotypes not identified among ABCs isolates tested for nonsusceptibility), we imputed this value as the inverse variance-weighted mean proportion of nonsusceptible isolates across serotypes with known susceptibility, using the Metafor package for R (v 4.6-0).^31^

## RESULTS

We estimated that US adults experience 58.1 (95% confidence interval [CI): 48.9-69.0) outpatient visits for non-bacteremic pneumococcal pneumonia, 380.9 (233.9-543.9) outpatient visits for pneumococcal sinusitis, and 2.5 (2.1-2.9) non-bacteremic pneumococcal pneumonia hospitalizations per 10,000 person-years (**Table 1**).

**Table 1.** Estimated incidence and annual numbers of pneumococcal pneumonia and sinusitis.

| Pneumococcal syndrome | Age group | Outpatient visits |  | Hospitalizations |  |
| --- | --- | --- | --- | --- | --- |
|  |  | *Incidence per 10,000 person-years (95% CI)<sup>a</sup>* | *Annual no. in thousands (95% CI)<sup>b</sup>* | *Incidence per 10,000 person-years (95% CI)<sup>c</sup>* | *Annual no. in thousands (95% CI)<sup>d</sup>* |
| Non-bacteremic pneumonia | 18-49 years | 14.0 (9.0, 20.7) | 193.5 (124.7, 286.0) | 0.3 (0.2, 0.5) | 4.6 (2.9, 6.6) |
|  | 50-64 years | 70.3 (52.6, 93.1) | 442.1 (330.8, 585.8) | 2.6 (2.0, 3.2) | 16.3 (12.9, 20.1) |
|  | ≥65 years<sup>e</sup> | 155.4 (121.6, 198.1) | 839.5 (657.1, 1,070.4) | 7.9 (6.4, 9.6) | 42.8 (34.5, 51.8) |
|  | All adults<sup>f</sup> | 58.1 (48.9, 69.0) | 1,483.1 (1,248.1, 1,760.7) | 2.5 (2.1, 2.9) | 63.8 (54.6, 73.7) |
| Sinusitis | 18-49 years | 357.8 (216.2, 524.8) | 4,949.1 (2,990.7, 7,257.9) | -- | -- |
|  | 50-64 years | 411.1 (245.7, 613.9) | 2,585.8 (1,545.3, 3,861.7) | -- | -- |
|  | ≥65 years<sup>e</sup> | 395.2 (234.5, 596.9) | 2,135.7 (1,267.2, 3,225.7) | -- | -- |
|  | All adults<sup>f</sup> | 380.9 (233.9, 543.9) | 9,722.0 (5,969.6, 13,883.0) | -- | -- |

Abbreviations: CI – confidence interval; No. -- number
<sup>a</sup> Estimated as all-cause pneumonia and sinusitis outpatient visits incidence and counts multiplied by previously-published estimates of pneumococcal attributable fractions in pneumonia<sup>3</sup> and sinusitis.<sup>4</sup> All-cause outpatient visits estimated using data from the National Ambulatory Medical Care Survey and National Hospital Ambulatory Medical Care Survey (NAMCS/NHAMCS) and MarketScan databases.
<sup>b</sup> Estimated by multiplying incidence estimates by 2019 Bridged-Race Census<sup>24</sup> estimates.
<sup>c</sup> Estimated by dividing annual burden estimates by 2019 Bridged-Race Census<sup>24</sup> estimates.
<sup>d</sup> Estimated using data from the National Inpatient Sample and Active Bacterial Core Surveillance, as detailed in **Table S2**.
<sup>e</sup> Estimates for outpatient settings derived from MarketScan (all outpatient settings except emergency departments [ED] and physician offices) imputed based on ratio of visits to ED and physician office visits to all other outpatient settings among adults aged 50-64 years multiplied by the incidence of physician office and ED visits observed in adults ≥65 years.
<sup>f</sup> Age-group annual no. values may not sum exactly to all adults totals due to uncertainty propagation methods and rounding.

The most prevalent serotypes were 3 (11.6% [8.7-15.0%]), 22F (7.8% [5.4-10.7%]), 20B (5.7% [3.6-8.3%]), 19A (5.6% [3.5-8.1%]), 35B (4.7% [2.8-7.2%]), and 9N (4.5% [2.7-6.9%]) (**Figure 1; Table S4**). Isolates from serotypes 15D, 6D, 15F, and 35A were universally nonsusceptible to ≥1 outpatient first-line antibiotic for pneumonia. Other serotypes with high prevalences of nonsusceptibility to ≥1 pneumonia outpatient first-line antibiotic included 35B (93.3% [90.8-95.3%]), 33F (92.5% [89.6-94.8%]), and 35D (89.2% [67.3-98.5]) (**Figure 2)**. Serotypes for which the greatest proportions of isolates were nonsusceptible to inpatient first-line antibiotics for pneumonia included 45 (100.0% [100.0-100.0%]), 35B (92.2% [89.6-94.4%]), 9V and 35D (both 89.2% [67.3-98.5%]), and 14 (78.4% [50.5-95.0%]). Serotypes with the greatest proportions of isolates nonsusceptible to the first-line agent for sinusitis (amoxicillin) included 35B (89.5% [86.5-92.0%]), 35D (89.2% [67.3-98.5%]), and 19A (48.8% [43.5-54.2%]).

**Figure 1.**
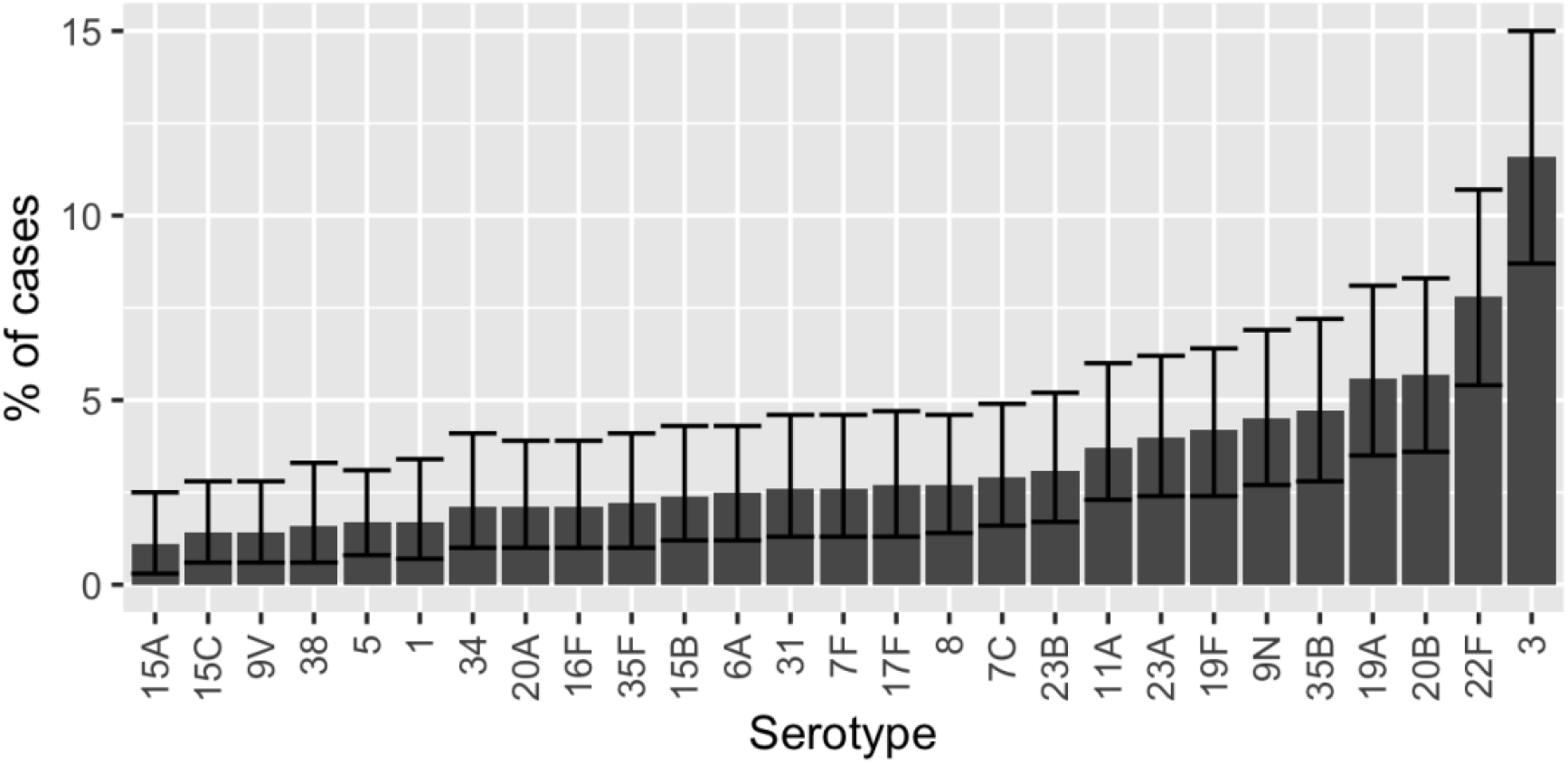
Serotype distribution in pneumococcal non-bacteremic pneumonia and sinusitis in adults. Only serotypes accounting for ≥1% of cases shown.

**Figure 2.**
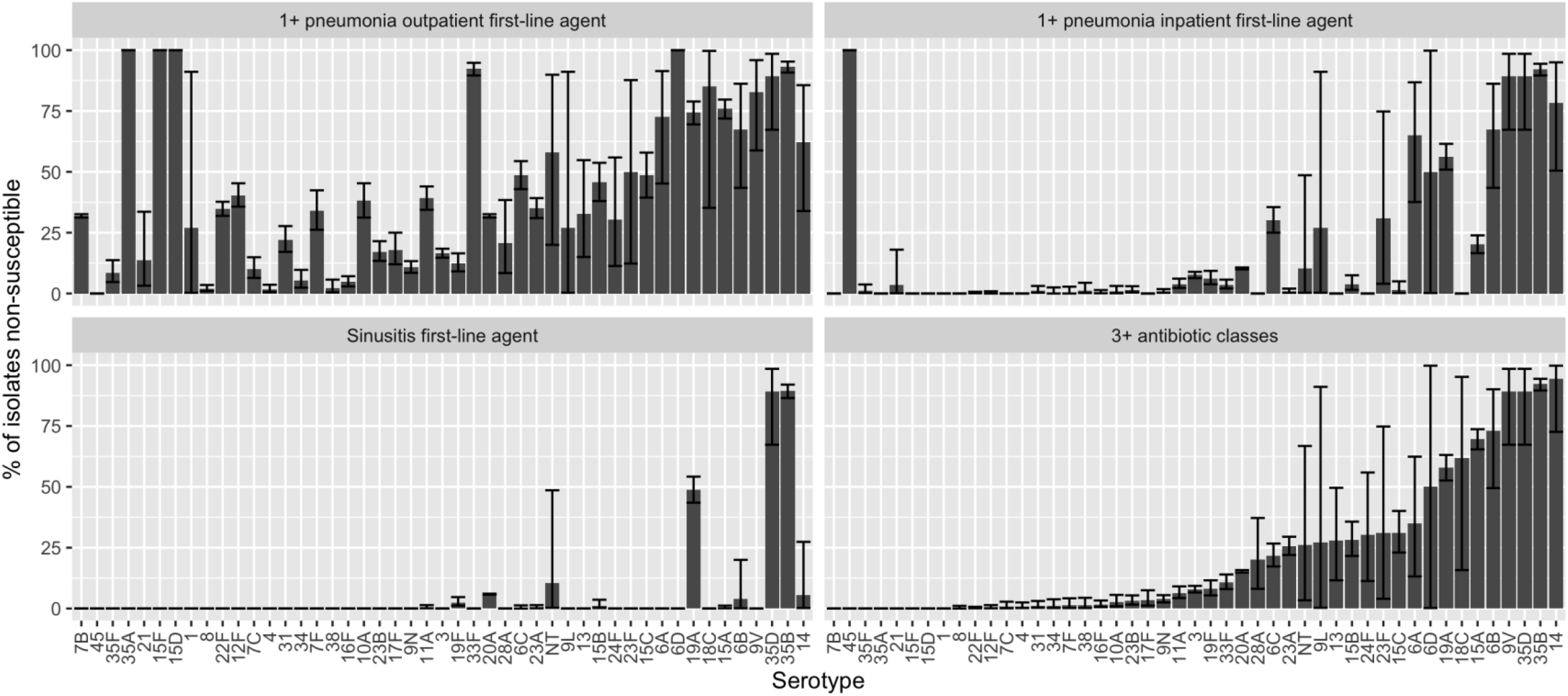
Percent of isolates nonsusceptible to first-line recommended agents and ≥3 antibiotic classes by serotype. Only serotypes with known (non-imputed) nonsusceptibility data from Active Bacterial Core surveillance and those with >1% of isolates nonsusceptible for any category shown. Percent of isolates nonsusceptible to 1 first-line agent for pneumonia imputed for serotype 7B (all other categories not imputed). Outpatient first-line antibiotic agents for adult pneumonia defined as amoxicillin, doxycycline, and erythromycin (proxy for all macrolides).^9^ Inpatient first-line antibiotic agents for adult pneumonia defined as cephalosporins (cefotaxime, ceftriaxone, and cefuroxime) and levofloxacin (proxy for all respiratory fluoroquinolones).^9^ First-line antibiotic agent for adult sinusitis defined as amoxicillin.^10,11^

Nonsusceptibility to ≥3 antibiotic classes was most common in serotypes 14 (94.5% [72.6-99.8%]), 35B (92.2% [89.6-94.4%]), 35D (89.2 [67.3-98.5%]), 9V (89.2% [67.3-98.5%]), and 6B (73.1% [49.5-90.1%]). Isolates were universally susceptible to linezolids and glycopeptides. Class and serotype-specific nonsusceptibility patterns are presented in **Table S4**.

Almost one-third (31.2% [26.6-36.3%]) of non-bacteremic pneumococcal pneumonia cases were expected to be nonsusceptible to ≥1 outpatient first-line antibiotic agent, translating to 462,206 (366,292-581,045) outpatient visits annually, and 13.7% (10.7-17.9%) were nonsusceptible to ≥2 outpatient first-line antibiotics (**Table 2**). An estimated 13.4% (9.8-18.5%), or 8,452 (5,925-12,077) pneumococcal pneumonia hospitalizations annually, were nonsusceptible to ≥1 inpatient first-line antibiotic. In total, 16.4% (12.8-21.4%) of pneumococcal pneumonia cases were nonsusceptible to ≥3 antibiotic classes, translating to 243,521 (179,673-333,675) outpatient visits and 10,155 (7,542-13,803) hospitalizations annually. We estimated that among non-bacteremic pneumococcal pneumonia cases, 28.0% (23.7-32.6%) were nonsusceptible to macrolides, 21.0% (17.2-25.7%) to antifolates, and 13.4% (9.7-18.5%) to cephalosporins, while only 0.1% (0.1-0.2%) were nonsusceptible to fluoroquinolones.

**Table 2.** Proportion and burdens of antibiotic nonsusceptible non-bacteremic pneumococcal pneumonia.

| Nonsusceptible to<sup>a</sup> | Percent of cases (95% CI)<sup>b</sup> | Outpatient visits |  | Hospitalizations |  |
| --- | --- | --- | --- | --- | --- |
|  |  | *Incidence per 10,000 person-years (95% CI)* | *Annual no. in thousands (95% CI)* | *Incidence per 10,000 person-years (95% CI)* | *Annual no. in thousands (95% CI)* |
| ≥1 outpatient first-line antibiotic agent<sup>c</sup> | 31.2 (26.6, 36.3) | 18.1 (14.1, 22.8) | 462.2 (366.3, 581.0) | 0.8 (0.6, 1.0) | 19.7 (15.8, 24.4) |
| ≥2 outpatient first-line antibiotic agents<sup>c</sup> | 13.7 (10.7, 17.9) | 8.0 (5.9, 10.9) | 203.7 (150.6, 278.3) | 0.3 (0.2, 0.5) | 8.5 (6.3, 11.5) |
| ≥1 inpatient first-line antibiotic agent<sup>d</sup> | 13.4 (9.8, 18.5) | 7.8 (5.4, 11.2) | 199.1 (139.0, 286.5) | 0.3 (0.2, 0.5) | 8.5 (5.9, 12.1) |
| >1 antibiotic class | 21.6 (17.3, 27.2) | 12.6 (9.5, 16.7) | 320.5 (242.4, 426.0) | 0.5 (0.4, 0.7) | 13.5 (10.3, 17.8) |
| ≥3 antibiotic classes | 16.4 (12.8, 21.4) | 9.5 (7.0, 13.1) | 243.5 (179.7, 333.7) | 0.4 (0.3, 0.5) | 10.2 (7.5, 13.8) |
| Macrolides | 28.0 (23.7, 32.6) | 16.3 (12.8, 20.5) | 415.0 (327.4, 522.3) | 0.7 (0.6, 0.9) | 17.7 (14.1, 21.9) |
| Antifolates | 21.0 (17.2, 25.7) | 12.2 (9.4, 15.9) | 311.5 (239.1, 406.8) | 0.5 (0.4, 0.7) | 13.4 (10.4, 17.3) |
| Cephalosporins | 13.4 (9.7, 18.5) | 7.8 (5.4, 11.2) | 198.3 (138.2, 285.6) | 0.3 (0.2, 0.5) | 8.4 (5.9, 12.0) |
| Tetracyclines | 12.8 (10.2, 16.4) | 7.4 (5.6, 10.0) | 189.7 (143.1, 255.9) | 0.3 (0.2, 0.4) | 8.0 (6.1, 10.8) |
| Carbapenems | 9.9 (6.5, 14.8) | 5.7 (3.7, 8.9) | 146.3 (93.9, 226.8) | 0.2 (0.2, 0.4) | 6.1 (3.9, 9.4) |
| Penicillins | 8.6 (5.7, 13.1) | 5.0 (3.2, 7.8) | 127.9 (81.7, 199.7) | 0.2 (0.1, 0.3) | 5.2 (3.3, 8.1) |
| Lincosamides | 8.6 (6.5, 11.8) | 5.0 (3.6, 7.1) | 127.3 (92.6, 182.4) | 0.2 (0.2, 0.3) | 5.3 (3.9, 7.5) |
| Fluoroquinolones | 0.1 (0.1, 0.2) | 0.1 (0.0, 0.1) | 1.8 (0.9, 3.7) | 0.0 (0.0, 0.0) | 0.1 (0.0, 0.1) |
| Linezolid<sup>e</sup> | 0.0 (0.0, 0.0) | 0.0 (0.0, 0.0) | 0 (0, 0) | 0.0 (0.0, 0.0) | 0 (0, 0) |
| Glycopeptides<sup>e</sup> | 0.0 (0.0, 0.0) | 0.0 (0.0, 0.0) | 0 (0, 0) | 0.0 (0.0, 0.0) | 0 (0, 0) |

Abbreviations: CI – confidence interval; No. – number
<sup>a</sup> Nonsusceptibility to antibiotic classes evaluated based on nonsusceptibility to specific agents within each class, as defined in **Table S2**.<sup>b</sup> Estimated as annual no. nonsusceptible pneumococcal outpatient visits for each nonsusceptibility category divided by annual no. total pneumococcal-attributable outpatient visits (N = 1,483,052 [95% CI 1,248,097, 1,760,694]).<sup>c</sup> Outpatient first-line antibiotic agents for adult pneumonia defined as amoxicillin, doxycycline, and erythromycin (proxy for all macrolides).<sup>9</sup><sup>d</sup> Inpatient first-line antibiotic agents for adult pneumonia defined as cephalosporins (cefotaxime, ceftriaxone, and cefuroxime) and levofloxacin (proxy for all respiratory fluoroquinolones).<sup>9</sup><sup>e</sup> All pneumococcal isolates captured in ABCs were susceptible to linezolid and glycopeptides.

We estimated that 8.7% (5.1-14.1%) and 9.7% (7.4-13.2%) of pneumococcal sinusitis cases were nonsusceptible to recommended first-line and alternative antibiotics, respectively, translating to 833,697 (414,093-1,536,416) and 942,808 (540,819-1,519,744) outpatient visits each year (**Table 3**). An estimated 19.0% (14.8-24.9%) of pneumococcal sinusitis cases were nonsusceptible to ≥3 antibiotic classes, translating to 1,844,726 (1,070,763-2,904,089) outpatient visits annually. Nonsusceptibility in pneumococcal sinusitis was most common for macrolides (29.7% [25.1-34.8]), antifolates (21.9% [17.9-26.6%]), and cephalosporins (15.2% [10.9-21.1%]).

**Table 3.** Proportion and burdens of antibiotic nonsusceptible pneumococcal sinusitis.

| Nonsusceptible to<sup>a</sup> | Percent of cases (95% CI)<sup>b</sup> | Outpatient visits |  |
| --- | --- | --- | --- |
|  |  | *Incidence per 10,000 person-years (95% CI)* | *Annual no. in thousands (95% CI)* |
| First-line antibiotic agent<sup>c</sup> | 8.7 (5.1, 14.1) | 32.7 (16.2, 60.2) | 833.7 (414.1, 1,536.4) |
| ≥1 alternative antibiotic agent<sup>d</sup> | 9.7 (7.4, 13.2) | 36.9 (21.2, 59.5) | 942.8 (540.8, 1,519.7) |
| >1 antibiotic class | 23.9 (19.3, 30.1) | 90.9 (53.6, 139.5) | 2,321.4 (1,368.8, 3,561.4) |
| ≥3 antibiotic classes | 19.0 (14.8, 24.9) | 72.3 (42.0, 113.8) | 1,844.7 (1,070.8, 2,904.1) |
| Macrolides | 29.7 (25.1, 34.8) | 112.9 (67.7, 167.6) | 2,881.3 (1,728.8, 4,277.0) |
| Antifolates | 21.9 (17.9, 26.6) | 83.2 (49.4, 125.8) | 2,122.7 (1,259.7, 3,212.0) |
| Cephalosporins | 15.2 (10.9, 21.1) | 57.4 (32.3, 94.1) | 1,466.2 (823.3, 2,400.9) |
| Tetracyclines | 13.5 (10.8, 17.0) | 51.2 (30.1, 78.8) | 1,308.0 (769.3, 2,011.8) |
| Carbapenems | 12.1 (8.0, 18.0) | 45.7 (24.5, 78.7) | 1,165.5 (624.6, 2,009.5) |
| Penicillins | 11.3 (7.7, 16.7) | 42.7 (23.3, 73.2) | 1,090.7 (593.8, 1,867.3) |
| Lincosamides | 9.6 (7.3, 13.1) | 36.5 (20.9, 58.9) | 930.4 (534.4, 1,502.7) |
| Fluoroquinolones | 0.2 (0.1, 0.4) | 0.6 (0.2, 1.5) | 15.9 (6.2, 39.0) |
| Linezolid<sup>e</sup> | 0.0 (0.0, 0.0) | 0.0 (0.0, 0.0) | 0 (0, 0) |
| Glycopeptides<sup>e</sup> | 0.0 (0.0, 0.0) | 0.0 (0.0, 0.0) | 0 (0, 0) |

Abbreviations: CI – confidence interval; No. – number
<sup>a</sup> Nonsusceptibility to antibiotic classes evaluated based on nonsusceptibility to specific agents within each class, as defined in **Table S2**.
<sup>b</sup> Estimated as annual no. nonsusceptible pneumococcal outpatient visits for each nonsusceptibility category divided by annual no. total pneumococcal-attributable outpatient visits (N = 9,722,028 [95% CI 5,969,625, 13,883,036]).
<sup>c</sup> First-line antibiotic agent for adult sinusitis defined as amoxicillin as a proxy for amoxicillin-clavulanate.<sup>10,11</sup>
<sup>d</sup> Alternative antibiotic agents for adult sinusitis defined as doxycycline, levofloxacin, clindamycin, and cephalosporins.<sup>10,11</sup>
<sup>e</sup> All pneumococcal isolates captured in ABCs were susceptible to linezolid and glycopeptides.

Considering both outpatient-managed pneumococcal sinusitis and non-bacteremic pneumonia together, an estimated 1,298,057 (836,840-2,050,628) ARIs were nonsusceptible to ≥1 outpatient first-line antibiotic agent annually. In total, 81.9 (50.5-125.1) outpatient-managed ARIs per 10,000 person-years, or 2,089,275 (1,289,201-3,193,219) ARIs annually, were nonsusceptible to ≥3 antibiotic classes.

The proportion of antibiotic-nonsusceptible cases of pneumococcal pneumonia and sinusitis were highest among adults aged 18-49 years compared with older age groups for all categories and classes (**Table S5-Table S8**). For instance, pneumococci nonsusceptible to ≥3 antibiotic classes accounted for 22.1% (16.6-29.5%) of non-bacteremic pneumococcal pneumonia and sinusitis among adults 18-49 years but 15.1% (11.2-20.4%) among those ≥65 years. However, given the higher overall incidence of pneumonia in older adults, the highest incidence rates and annual burdens of antibiotic nonsusceptible non-bacteremic pneumococcal pneumonia were observed among adults aged ≥65 years (**Table S5, Table S6**). In pneumococcal sinusitis, where younger age groups account for higher incidence rates and annual case numbers compared with adults aged ≥65 years (**Table 1**), the greatest incidence of nonsusceptible cases occurred among adults aged 18-49 years for most categories and classes (**Table S7, Table S8**).

For non-bacteremic pneumococcal pneumonia, cases attributable to serotypes targeted by PCV15, PCV20, and PCV21 that were nonsusceptible to ≥1 outpatient first-line antibiotic accounted for 16.0% (12.1-21.0%), 19.2% (15.3-23.9%), and 24.8% (20.3-30.1%) of cases, respectively, while similarly nonsusceptible NVT serotypes accounted for only 1.8% (0.9-5.1%) of cases (**Table 4**). PCV15-serotypes nonsusceptible to ≥1 inpatient first-line antibiotic accounted for 8.0% (5.1-11.8%) of non-bacteremic pneumococcal pneumonia cases while PCV20 and PCV21 serotypes nonsusceptible to ≥1 inpatient first-line antibiotic were responsible for 8.3% (5.4-12.1%) and 10.8% (7.3-15.9%) of cases, respectively. Serotypes targeted by PCV15, PCV20, and PCV21 with nonsusceptibility to ≥3 antibiotic classes accounted for 7.4% (4.7-11.1%), 8.5% (5.8-12.1%), and 12.6% (9.2-17.5%) of non-bacteremic pneumococcal pneumonia cases, respectively. Nonsusceptible NVT serotypes contributed minimally across all measures. Divergent trends were observed by antibiotic class (**Table S9**).

**Table 4.**
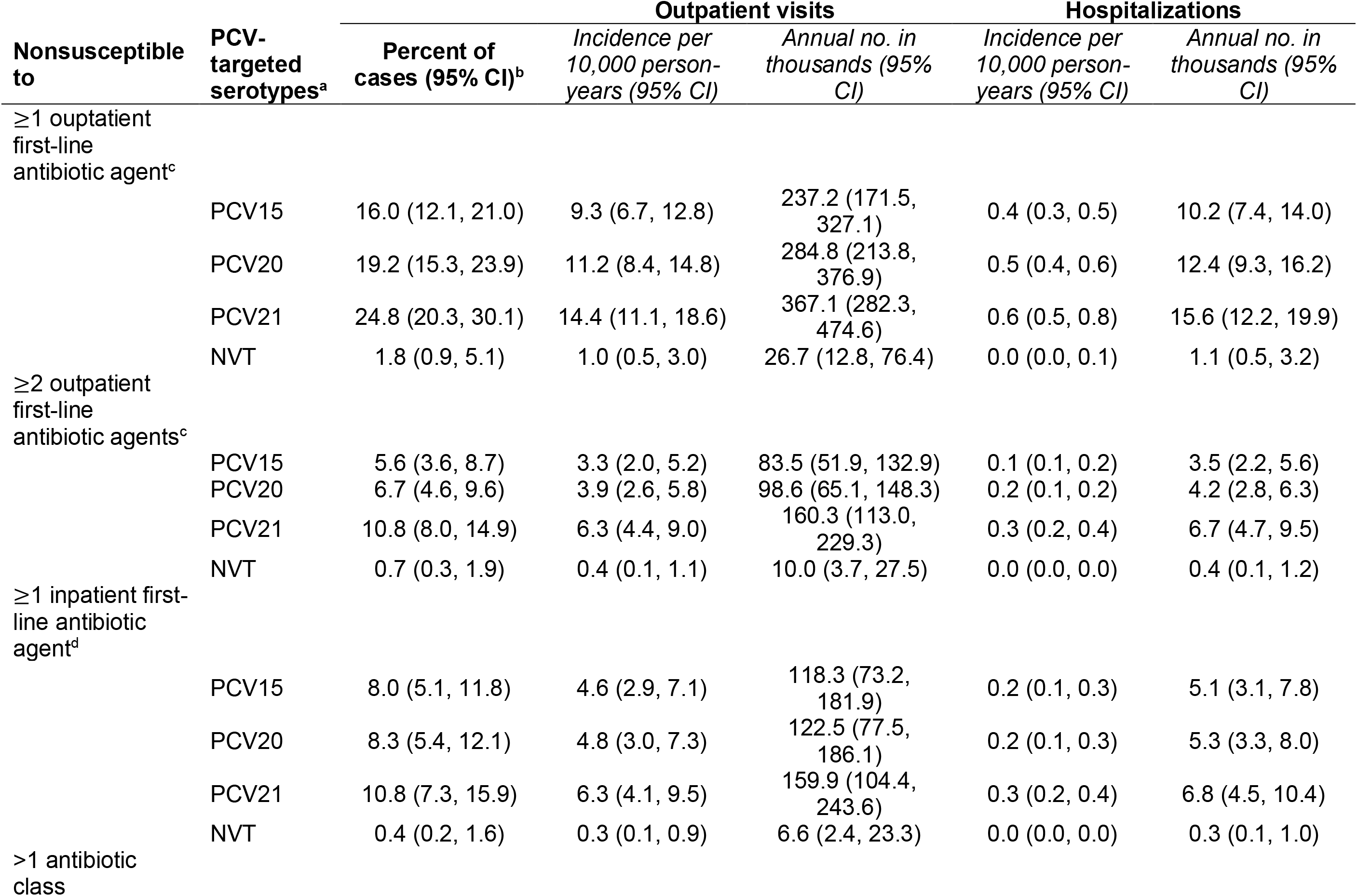
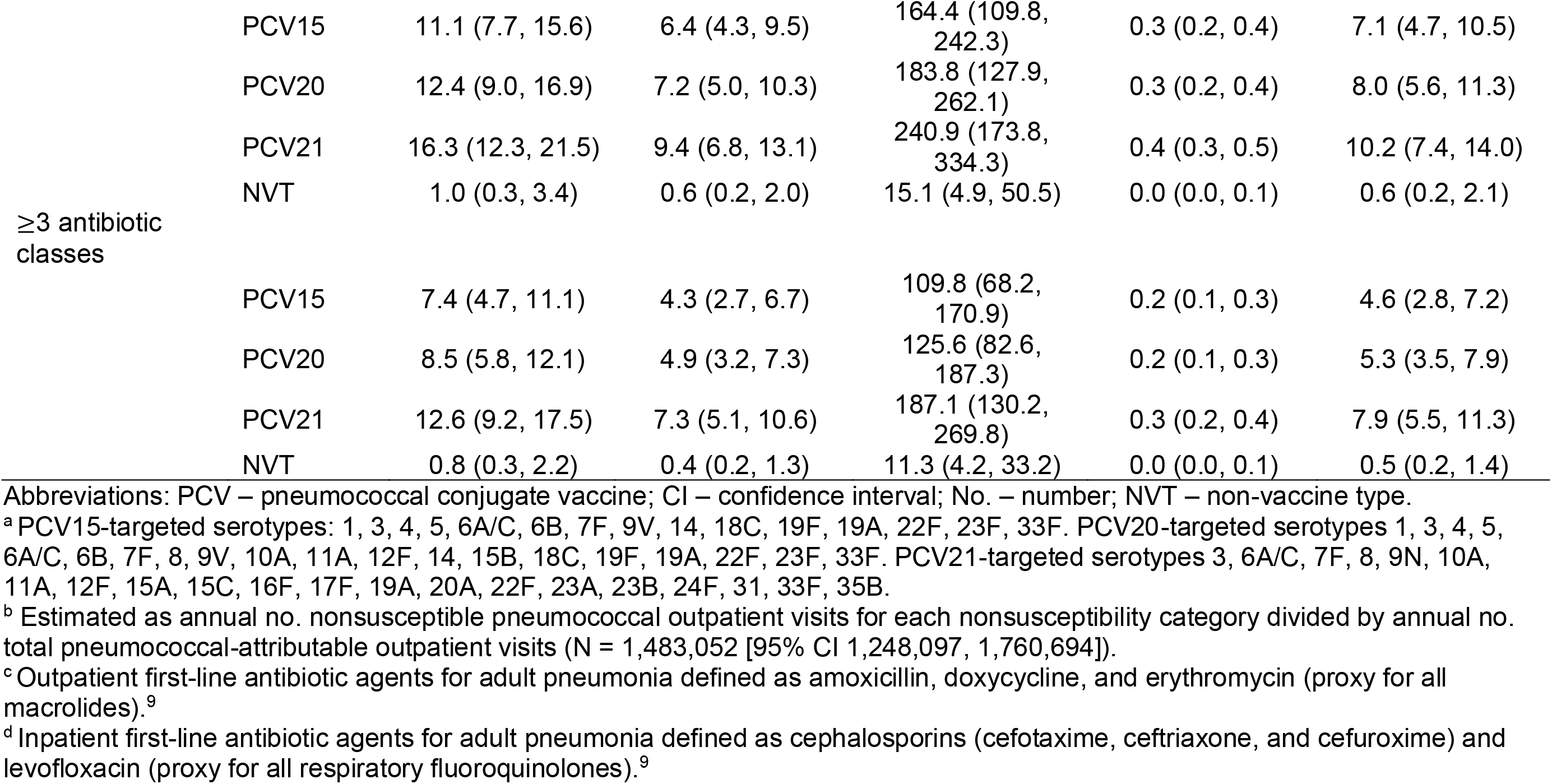
Burdens of antibiotic-nonsusceptible non-bacteremic pneumococcal pneumonia by PCV-targeted serotypes.

Antibiotic nonsusceptible PCV-targeted serotypes accounted for smaller proportions of pneumococcal sinusitis compared with pneumonia (**Table 5**). Serotypes targeted by PCV15, PCV20, and PCV21 that were nonsusceptible to the first-line antibiotic accounted for 3.0% (1.4-5.6%), 3.1% (1.4-5.7%), and 8.3% (4.7-13.8%) of pneumococcal sinusitis cases. As in pneumonia, NVT serotypes contributed minimally to antibiotic-nonsusceptible sinusitis burdens (**Table 5, Table S10**). For all classes except antifolates, PCV21 serotype burdens were greater than those due to PCV15, PCV20, and NVT serotypes (**Table S10**).

**Table 5.**
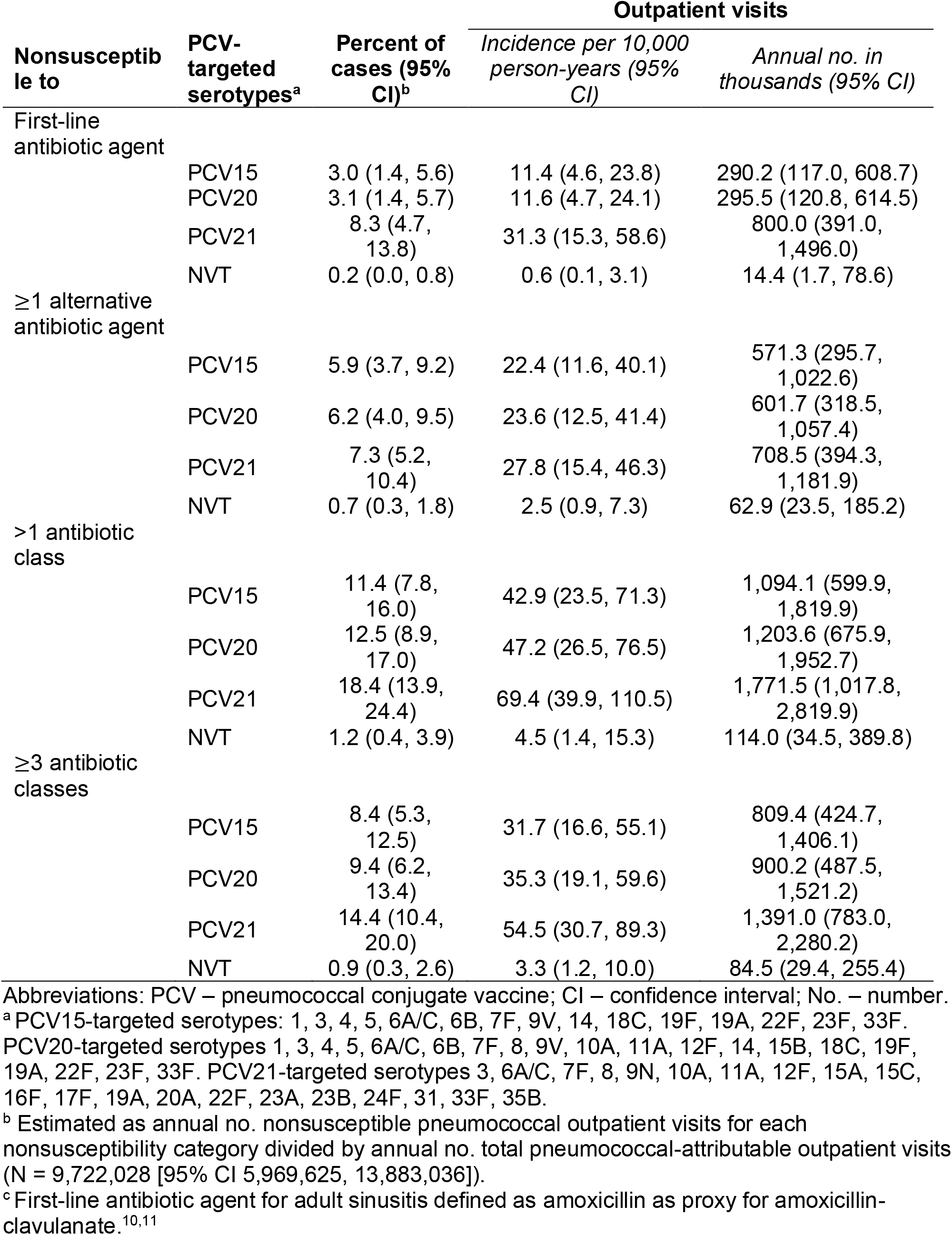
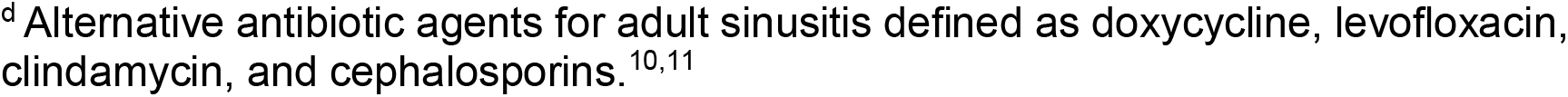
Burdens of antibiotic-nonsusceptible pneumococcal sinusitis by PCV-targeted serotypes.

In sensitivity analyses applying serotype distribution from pediatric ARIs to adult sinusitis, the most prevalent serotypes were 15C (11.4% [10.5-12.2%]), 23B (9.3% [8.6-10.0%]), 11A (8.8% [8.2-9.6%]), 15A (7.8% [7.1-8.4%]), and 35B (7.6% [6.9-8.2%]). Compared with primary analyses, the incidence of pneumococcal sinusitis nonsusceptible to the first-line antibiotic increased to 40.1 (24.3-58.6) cases per 10,000 person years and sinusitis cases nonsusceptible to ≥3 antibiotic classes increased to 104.1 (63.6-150.1) cases per 10,000 person years (**Table S11**). Additionally, antibiotic-nonsusceptible NVT serotypes accounted for greater proportions of pneumococcal sinusitis cases compared with primary estimates (**Table S12**).

## DISCUSSION

We estimated that 16% of adult non-bacteremic pneumococcal pneumonia cases and 19% of pneumococcal sinusitis cases were multidrug resistant, i.e., nonsusceptible to ≥3 antibiotic classes. These proportions translate to 244 thousand outpatient visits and 10 thousand hospitalizations for multidrug-resistant non-bacteremic pneumonia and 1.8 million outpatient visits for multidrug-resistant sinusitis. An estimated 31% of non-bacteremic pneumococcal pneumonia cases were nonsusceptible to ≥1 outpatient first-line antibiotic while only 14% of cases were nonsusceptible to ≥2 outpatient first-line agents and 13% were nonsusceptible to ≥1 inpatient first-line agent. For sinusitis, only 9% of cases were nonsusceptible to the first-line agent. Thus, most cases of pneumococcal ARIs among adults in the United States can still be effectively managed with first-line or alternative agents, although initial treatment failure, which can lead to higher costs and more intensive care, is possible, especially in outpatient-managed pneumonia.

Across most nonsusceptibility measures, PCV21-targeted serotypes accounted for greater proportions of pneumococcal non-bacteremic pneumonia and sinusitis cases than PCV15- and PCV20-targeted serotypes. Notably, NVT serotypes accounted for small proportions of nonsusceptible pneumococcal pneumonia: NVT serotypes nonsusceptible to first-line treatments for pneumonia accounted for 1.8% of outpatient visits and 0.4% of hospitalizations for non-bacteremic pneumococcal pneumonia. Similar findings were observed for sinusitis in primary analyses. Thus, current PCVs target serotypes responsible for antibiotic nonsusceptible infections and may be powerful tools to mitigate antibiotic nonsusceptible pneumococcal disease. Historically, implementation of PCV7 and PCV13 resulted in changes in serotype distribution^32^ and reductions in vaccine-serotype antibiotic-nonsusceptible pneumococcal infection burdens, with some decreases offset by increases in NVT antibiotic-nonsusceptible disease.^13,17,29^

Certain PCV-targeted serotypes contributed meaningfully to overall nonsusceptibility burdens. Serotype 35B, prevalent in both pneumococcal sinusitis and pneumonia, was nonsusceptible to almost all drug classes and 92.2% of 35B isolates were nonsusceptible to ≥3 agents. PCV21 is the only currently-licensed pneumococcal vaccine targeting serotype 35B. Pipeline PCVs also target this serotype.^27^ Serotype 19A—which is prevalent in adult pneumococcal ARIs and for which a majority of isolates are also nonsusceptible to most classes—was targeted by PCV13 and is included in the formulation of all PCVs currently recommended for use in adults. Its persistence in adult disease may reflect low PCV uptake in this group; in 2021, only 66% of US adults aged ≥65 years universally eligible for PCV vaccination and 22% of US adults 19-64 years eligible for risk-based PCV vaccination had ever received pneumococcal vaccination (including both PCVs and the 23-valent pneumococcal polysaccharide vaccine).^33^ Further uptake of PCVs in adults, potentially aided by the October 2024 expansion of universal vaccination recommendations to adults aged 50-64 years,^20^ may contribute to reducing the prevalence of this highly nonsusceptible serotype. Serotype 3, previously targeted by PCV13 and now targeted by PCV15/20/21, still accounted for the highest proportion (12%) of all serotypes in pneumococcal pneumonia. Despite low overall rates of nonsusceptibility in serotype 3, due to its overall prevalence, it wields outsize influence on susceptibility distributions in pneumococci at large. Ongoing surveillance of serotype distribution in non-invasive disease and antibiotic nonsusceptibility in non-vaccine-type serotypes will be important in informing future PCV formulations.

Antibiotic resistance is driven by antibiotic consumption, either for infections caused by *S. pneumoniae* or due to exposure of pneumococcus to antibiotics for other infections.^34^ Thus, improving antibiotic prescribing practices and reducing antibiotic use through infection prevention remain important objectives. We found that antibiotic nonsusceptibility in pneumococcal ARIs was highest for macrolides compared with all other antibiotic classes. Macrolides are commonly prescribed for conditions for which they are not recommended (e.g., sinusitis, AOM) and those for which no antibiotics are recommended (e.g., viral upper respiratory infection, bronchitis).^4,35,36^ A modeling study estimated that 93% of pneumococcal macrolide exposures occurred when antibiotics were consumed for infections caused by non-pneumococcal pathogens.^34^ We also observed prevalent nonsusceptibility to antifolates in pneumonia. Antifolates are not indicated in adults for ARIs commonly caused by *S. pneumoniae* but are recommended in the management of urinary tract infections (UTIs),^37,38^ a leading cause of antibiotic use among adults.^39,40^ We observed low proportions of fluoroquinolone nonsusceptibility in pneumococcal ARIs despite fluoroquinolones being frequently prescribed in adults: in 2013-14 40% of genitourinary and 15% of respiratory infections in adult outpatients were treated with fluoroquinolones.^41^ Lack of fluoroquinolone resistance may be related to fitness tradeoffs for fluoroquinolone-specific resistance mechanisms^42^ and the fact that young children, important in pneumococcal colonization and transmission,^43,44^ are infrequently prescribed fluoroquinolones.^45^

Our study has limitations. First, to capture nonsusceptibility proportions for as many serotypes as possible, we used data from IPD isolates. Nonsusceptibility patterns in non-invasive disease may vary from those in invasive disease, although there is limited understanding of potential serotype-specific nonsusceptibility differences in adults. A study in children in Rochester, NY found similar serotype-specific antibiotic nonsusceptibility patterns between AOM and IPD isolates for penicillins and macrolides but divergent patterns for cephalosporins and tetracyclines.^16^ A meta-analysis of studies of antibiotic nonsusceptibility in *S. pneumoniae* (adult and pediatric) found similar rates of nonsusceptibility to penicillin and macrolides in pneumococcal carriage and invasive disease isolates in high-income countries.^17^ Overall nonsusceptibility was similar or lower in isolates from invasive sources compared with non-invasive sources in a study of U.S. adults with pneumonia hospitalized at SENTRY Antimicrobial Surveillance Program sites from 2009-2017, however serotype distribution was not considered.^46^ For serotypes not identified in ABCs isolates, we imputed nonsusceptibility based on weighted averages for all identified serotypes. As serotypes not contained in ABCs data accounted for few ARI cases this assumption is unlikely to have meaningfully affected our overall estimates. Second, serotype distribution was based on data from hospitalized adults in the Southeastern United States^3^ and may not be representative of all US adults and those treated in outpatient settings. We supplemented this data with IPD serotype distribution, which may not reflect serotype distribution in non-bacteremic pneumonia. Third, data on post-PCV13 sinusitis etiology are limited. We relied on serotype distribution in adult pneumonia, supplemented with sensitivity analyses using serotype distribution from pediatric ARIs (AOM and pneumonia). In sensitivity analyses, NVT serotypes contributed markedly to antibiotic-nonsusceptible sinusitis burdens, emphasizing the importance of etiologic assumptions. Fourth, we did not consider serotype cross-protection in estimating PCV-serotype-attributable burdens. Thus, our estimates of PCV-associated burdens are likely underestimates. Finally, burden estimates were based on national surveys and administrative datasets and relied on ICD-10-CM codes to identify diagnoses.

In conclusion, we demonstrated high proportions of antibiotic nonsusceptibility in pneumococcal ARIs in US adults. We found that 16% of non-bacteremic pneumonia cases and 19% of pneumococcal sinusitis cases were nonsusceptible to ≥3 antibiotic classes. However, most cases were susceptible to at least one first-line or alternative recommended agent. Serotypes targeted by PCV15, PCV20, and PCV21 accounted for sizeable burdens of antibiotic-nonsusceptible cases compared with NVT serotypes. Uptake of these vaccines and ongoing antibiotic stewardship may contribute to addressing antibiotic nonsusceptibility in pneumococcal disease.

## FOOTNOTES

### Disclaimers

This work was supported by the Centers for Disease Control and Prevention [21IPA2111845 to JAL] and the National Institutes of Health [1F31AI174773 to LMK]. The National Institutes of Health had no input into the design and conduct of the study; collection, management, analysis, and interpretation of the data; preparation, review, or approval of the manuscript; and decision to submit the manuscript for publication. The Centers of Disease Control and Prevention was involved through co-author participation the design and conduct of the study; analysis and interpretation of the data; preparation, review, or approval of the manuscript; and decision to submit the manuscript for publication. The content is solely the responsibility of the authors and does not necessarily represent the official views of the National Institutes of Health and the Centers for Disease Control and Prevention.

### Data availability

NAMCS, NHAMCS and NIS datasets are publicly available. MarketScan data is proprietary. ABCs data is available by request to https://www.cdc.gov/abcs/isolates-and-data/isolate-and-data-requests.html.

### Author contributions

**Laura M King:** Conceptualization, methodology, validation, formal analysis, investigation, data curation, writing – original draft. **Kristin L Andrejko:** Conceptualization, resources, writing - review & editing, supervision, project administration. **Miwako Kobayashi:** conceptualization, methodology, resources, writing - review & editing. **Wei Xing:** Formal analysis, writing - review & editing. **Adam L Cohen:** Conceptualization, resources, writing - review & editing. **Joseph A Lewnard:** Conceptualization, methodology, writing - review & editing, supervision, resources, funding acquisition.

