## Supplemental materials for "Acute respiratory infections due to antibiotic-nonsusceptible *Streptococcus pneumoniae* in United States adults"

#### CONTENTS

**Table S1.** *International Classification of Diseases, 10th revision, Clinical Modification* (ICD-10-CM) codes used to identify pneumonia and sinusitis for all-cause incidence estimates

**Table S2.** Estimated all-cause, all pneumococcal, and bacteremic pneumococcal pneumonia cases, 2019

**Table S3.** Antibiotic classes and agents for which *Streptococcus pneumoniae* isolates were tested for nonsusceptibility in CDC's Active Bacterial Core surveillance

**Table S4.** Serotype distribution and percent of isolates nonsusceptible by drug class and serotype

**Table S5.** Percent and burdens of antibiotic-nonsusceptible non-bacteremic pneumococcal pneumonia by nonsusceptibility criteria and patient age group

**Table S6.** Percent and burdens of antibiotic-nonsusceptible non-bacteremic pneumococcal pneumonia by drug class and patient age group

**Table S7.** Percent and burdens of antibiotic-nonsusceptible pneumococcal sinusitis by nonsusceptibility criteria and patient age group

**Table S8.** Percent and burdens of antibiotic-nonsusceptible pneumococcal sinusitis by drug class and patient age group

**Table S9.** Burdens of antibiotic-nonsusceptible non-bacteremic pneumococcal pneumonia by antibiotic class and PCV-targeted serotypes

**Table S10.** Burdens of antibiotic-nonsusceptible pneumococcal sinusitis by antibiotic class and PCV-targeted serotypes

**Table S11.** Sensitivity analysis: Burdens of antibiotic nonsusceptible pneumococcal sinusitis assuming serotype distribution based on meta-analysis of studies of pediatric AOM and pneumonia

**Table S12.** Sensitivity analysis burdens of antibiotic-nonsusceptible pneumococcal sinusitis by PCV-targeted serotypes assuming serotype distribution based on meta-analysis of studies of pediatric AOM and pneumonia

### Supplemental Material References

**Table S1.** ***International Classification of Diseases, 10th revision, Clinical Modification*** **(ICD-10-CM) codes used to identify pneumonia and sinusitis for all-cause incidence estimates**

| **Condition** | **Setting[a]** | **ICD-10-CM codes** |
|---|---|---|
| Pneumonia | Outpatient | A48.1, B01.2, B05.2, B06.81,[b] J09.X1,[b] J10.0*, J11.0*, J10.0, J10.00, J10.01, J10.08, J11.0, J11.00, J11.08, J12, J12.0, J12.1, J12.2, J12.3, J12.8, J12.81, J12.82, J12.89, J12.9, J13, J14, J15, J15.0, J15.1, J15.2, J15.20, J15.21, J15.211, J15.212, J15.29, J15.3, J15..4, J15.5, J15.6, J15.7, J15.8, J15.9, J16, J16.0, J16.8, J17, J18, J18.0, J18.1, J18.2, J18.8, J18.9, J69, J690, J84.116,[b] J95.851,[b] O29.01,[b] O29.011,[b] O29.012,[b] O29.013,[b] O29.019,[b] O74.0, O89.01,[b] P23, P23.0, P23.1, P23.2, P23.3, P23.4, P23.5, P23.6, P23.8, P23.9 |
| Pneumonia | Inpatient | A01.03, A02.22, A15, A15.0, A15.7, A15.8, A15.9, A20.2, A21.2, A22.1, A37.01, A37.11, A37.81, A37.91, A42.0, A43.0, A48.1, A50.04, A54.84, B0.12, B05.2, B06.81, B2.50, B37.1, B38.0, B38.1, B38.2, B39.0, B39.1, B39.2, B44.0, B44.1, B58.3, B59, B77.81, J09.X1, J10.0, J10.00, J10.01, J10.08, J11.0, J11.00, J11.08, J12, J12.0, J12.1, J12.2, J12.3, J12.8, J12.81, J12.82, J12.89, J12.9, J13, J14, J15, J15.0, J15.1, J15.2, J15.20, J15.21, J15.211, J15.212, J15.29, J15.3, J15..4, J15.5, J15.6, J15.7, J15.8, J15.9, J16, J16.0, J16.8, J17, J18, J18.0, J18.1, J18.2, J18.8, J18.9, J85.1, P23, P23.0, P23.1, P23.2, P23.3, P23.4, P23.5, P23.6, P23.8, P23.9 |
| Sinusitis | Outpatient | J01*, J32* |

[a] Outpatient data from MarketScan databases and the National Ambulatory and National Hospital Ambulatory (NAMCS/NHAMCS) datasets. Inpatient data from the National Inpatient Sample (NIS).
[b] Only included in MarketScan incidence estimates due to truncation of ICD-10-CM codes in NAMCS/NHAMCS.

**Table S2. Estimated all-cause, all pneumococcal, and bacteremic pneumococcal pneumonia cases, 2019**

| | Estimated No. (95% CI) | | |
|---|---|---|---|
| **Age group** | *All-cause pneumonia hospitalizations*[a] | *Pneumococcal pneumonia hospitalizations*[b] | *Bacteremic pneumonia cases*[c] |
| 18-49 years | 90,155 (87,845, 92,507) | 8,966 (7,251, 10,905) | 4,352 |
| 50-64 years | 159,233 (155,403, 163,127) | 23,396 (19,941, 27,171) | 7,075 |
| ≥65 years | 457,544 (447,248, 467,986) | 51,968 (43,763, 61,032) | 9,215 |
| All adults[d] | 706,948 (695,704, 718,306) | 84,416 (75,244, 94,334) | 20,642 |

Abbreviations: CI – confidence interval

[a] Estimated as 2019 National Inpatient Sample (NIS)[1] estimates fit to gamma distributions.

[b] Estimated as all-cause pneumonia hospitalizations multiplied by pneumococcal attributable proportions from Self et al. 2024.[2]

[c] Estimated as 72.1%[3] of all invasive pneumococcal disease cases estimated using IPD incidence rates from 2019 Active Bacterial Core surveillance[3] (ABCs) projected to national estimates based on 2019 Bridged-Race Census estimates.[4] No uncertainty estimated consistent with original ABCs estimates.

[d] Sum of estimates from all age groups. Component age group values presented may not sum exactly to presented totals for all adults due to uncertainty propagation methods and rounding.

**Table S3. Antibiotic classes and agents for which *Streptococcus pneumoniae* isolates were tested for nonsusceptibility in CDC's Active Bacterial Core surveillance**

| **Class** | **Agent(s) tested for via Whole Genome Sequencing** |
|---|---|
| Penicillins | Penicillin, Amoxicillin |
| Macrolides | Erythromycin |
| Cephalosporins | Ceftriaxone, Cefuroxime, Cefotaxime |
| Tetracycline | Tetracycline, Doxycyline |
| Fluoroquinolones | Levofloxacin |
| Antifolates | Sulfamethoxazole-trimethoprim |
| Lincosamides | Clindamycin |
| Glycopeptides | Vancomycin |
| Carbapenems | Meropenem |
| Linezolids | Linezolid |

### Table S4. Serotype distribution and percent of isolates nonsusceptible by drug class by serotype

| Serotype | % of pneumococcal cases (95% CI)[a] | % of isolates nonsusceptible to antibiotic classes, all adults (95% CI)[b] | | | | | | | |
|---|---|---|---|---|---|---|---|---|---|
| | | *Cephalosporin* | *Lincosamide* | *Antifolate* | *Fluoroquino-lone* | *Carbapenem* | *Penicillin* | *Tetracycline* | *Macrolide* |
| 3 | 12 (9, 15) | 7 (6, 9) | 8 (7, 9) | 1 (0, 1) | 0 (0, 0) | 0 (0, 1) | 0 (0, 0) | 16 (14, 18) | 11 (9, 12) |
| 22F | 8 (5, 11) | 0 (0, 1) | 1 (0, 1) | 1 (1, 2) | 0 (0, 0) | 0 (0, 0) | 0 (0, 0) | 1 (0, 1) | 34 (31, 37) |
| 19A | 6 (4, 8) | 56 (51, 62) | 51 (46, 57) | 62 (56, 67) | 1 (0, 3) | 53 (48, 59) | 50 (45, 55) | 55 (49, 60) | 71 (66, 75) |
| 20B | 6 (4, 8) | 1 (0, 3) | 2 (1, 4) | 2 (1, 4) | 0 (0, 0) | 0 (0, 0) | 0 (0, 0) | 2 (1, 4) | 2 (1, 4) |
| 35B | 5 (3, 7) | 92 (90, 94) | 1 (0, 2) | 24 (20, 28) | 0 (0, 1) | 92 (90, 94) | 90 (87, 93) | 3 (2, 5) | 80 (77, 84) |
| 9N | 5 (3, 7) | 1 (0, 1) | 4 (2, 5) | 4 (3, 6) | 0 (0, 1) | 0 (0, 1) | 0 (0, 1) | 4 (3, 6) | 9 (7, 12) |
| 11A | 4 (2, 6) | 4 (2, 6) | 6 (4, 9) | 16 (13, 20) | 0 (0, 0) | 4 (2, 6) | 1 (1, 3) | 6 (4, 9) | 39 (34, 44) |
| 19F | 4 (2, 6) | 6 (4, 9) | 8 (6, 12) | 12 (9, 16) | 0 (0, 1) | 5 (3, 7) | 4 (2, 7) | 10 (7, 13) | 11 (8, 15) |
| 23A | 4 (2, 6) | 1 (0, 2) | 25 (22, 29) | 16 (13, 20) | 0 (0, 0) | 1 (0, 1) | 9 (6, 11) | 26 (22, 30) | 34 (30, 39) |
| 23B | 3 (2, 5) | 1 (0, 3) | 2 (1, 4) | 50 (45, 55) | 0 (0, 0) | 0 (0, 1) | 14 (11, 18) | 3 (1, 5) | 17 (13, 21) |
| 7C | 3 (2, 5) | 0 (0, 0) | 1 (0, 4) | 9 (5, 13) | 0 (0, 0) | 0 (0, 0) | 0 (0, 0) | 6 (3, 10) | 5 (2, 9) |
| 17F | 3 (1, 5) | 0 (0, 0) | 4 (1, 8) | 4 (1, 8) | 0 (0, 0) | 0 (0, 0) | 1 (0, 3) | 7 (4, 12) | 16 (11, 23) |
| 31 | 3 (1, 5) | 1 (0, 3) | 1 (0, 2) | 1 (0, 3) | 0 (0, 0) | 0 (0, 2) | 0 (0, 0) | 2 (1, 4) | 22 (17, 28) |
| 7F | 3 (1, 5) | 0 (0, 0) | 1 (0, 4) | 1 (0, 4) | 1 (0, 3) | 1 (0, 3) | 0 (0, 0) | 4 (1, 8) | 32 (24, 40) |
| 8 | 3 (1, 5) | 0 (0, 0) | 0 (0, 1) | 0 (0, 1) | 0 (0, 0) | 0 (0, 0) | 0 (0, 0) | 2 (1, 3) | 0 (0, 1) |
| 6A | 3 (1, 4) | 65 (38, 87) | 5 (0, 25) | 35 (13, 62) | 0 (0, 0) | 12 (2, 37) | 0 (0, 0) | 5 (0, 25) | 73 (45, 91) |
| 15B | 2 (1, 4) | 4 (1, 8) | 3 (1, 7) | 37 (30, 45) | 0 (0, 0) | 3 (1, 7) | 6 (3, 11) | 27 (20, 34) | 44 (37, 52) |
| 16F | 2 (1, 4) | 0 (0, 1) | 2 (1, 3) | 3 (2, 5) | 0 (0, 0) | 0 (0, 0) | 0 (0, 0) | 2 (1, 4) | 4 (3, 7) |
| 20A | 2 (1, 4) | 10 (10, 11) [c] | 9 (9, 10) [c] | 18 (17, 18) [c] | 0 (0, 0) [c] | 7 (7, 8) [c] | 8 (7, 8) [c] | 12 (11, 12) [c] | 30 (29, 30) [c] |
| 34 | 2 (1, 4) | 0 (0, 3) | 1 (0, 4) | 8 (4, 13) | 0 (0, 0) | 0 (0, 0) | 0 (0, 0) | 1 (0, 4) | 5 (2, 10) |
| 35F | 2 (1, 4) | 1 (0, 4) | 0 (0, 0) | 0 (0, 2) | 0 (0, 0) | 0 (0, 0) | 0 (0, 0) | 0 (0, 2) | 8 (5, 13) |
| 1 | 2 (1, 3) | 0 (0, 0) | 0 (0, 0) | 100 (100, 100) | 0 (0, 0) | 0 (0, 0) | 0 (0, 0) | 27 (0, 91) | 0 (0, 0) |
| 38 | 2 (1, 3) | 1 (0, 4) | 1 (0, 3) | 1 (0, 4) | 0 (0, 0) | 0 (0, 0) | 0 (0, 0) | 2 (0, 6) | 1 (0, 4) |
| 5 | 2 (1, 3) | 0 (0, 0) | 0 (0, 0) | 100 (100, 100) | 0 (0, 0) | 0 (0, 0) | 0 (0, 0) | 0 (0, 0) | 0 (0, 0) |
| 15C | 1 (1, 3) | 1 (0, 3) | 2 (0, 5) | 40 (31, 49) | 1 (0, 3) | 1 (0, 3) | 10 (5, 16) | 31 (23, 40) | 48 (39, 57) |
| 9V | 1 (1, 3) | 89 (67, 98) | 30 (11, 56) | 96 (78, 100) | 0 (0, 0) | 89 (67, 98) | 30 (11, 56) | 43 (21, 68) | 83 (59, 96) |

| Serotype | % of pneumococcal cases (95% CI)[a] | % of isolates nonsusceptible to antibiotic classes, all adults (95% CI)[b] | | | | | | | |
|---|---|---|---|---|---|---|---|---|---|
| | | *Cephalosporin* | *Lincosamide* | *Antifolate* | *Fluoroquino-lone* | *Carbapenem* | *Penicillin* | *Tetracycline* | *Macrolide* |
| 15A | 1 (0, 2) | 20 (17, 24) | 68 (64, 72) | 34 (29, 38) | 0 (0, 0) | 7 (5, 10) | 7 (5, 9) | 69 (65, 73) | 75 (71, 79) |
| 24F | 1 (0, 2) | 0 (0, 0) | 30 (11, 56) | 24 (7, 49) | 0 (0, 0) | 0 (0, 0) | 0 (0, 0) | 30 (11, 56) | 30 (11, 56) |
| 33F | 1 (0, 2) | 4 (2, 6) | 7 (5, 10) | 90 (86, 92) | 0 (0, 0) | 4 (2, 6) | 1 (0, 2) | 7 (5, 10) | 92 (89, 95) |
| 4 | 1 (0, 2) | 0 (0, 0) | 1 (0, 3) | 83 (78, 87) | 0 (0, 0) | 0 (0, 0) | 0 (0, 0) | 1 (0, 2) | 2 (1, 4) |
| 6B | 1 (0, 2) | 67 (43, 86) | 56 (32, 78) | 90 (71, 99) | 0 (0, 0) | 27 (10, 51) | 10 (1, 29) | 56 (32, 78) | 67 (43, 86) |
| 10A | 0 (0, 1) | 1 (0, 3) | 3 (1, 6) | 5 (2, 9) | 0 (0, 0) | 0 (0, 0) | 0 (0, 0) | 4 (2, 8) | 36 (29, 43) |
| 10D | 0 (0, 1) | 10 (10, 11) [c] | 9 (9, 10) [c] | 18 (17, 18) [c] | 0 (0, 0) [c] | 7 (7, 8) [c] | 8 (7, 8) [c] | 12 (11, 12) [c] | 30 (29, 30) [c] |
| 12F | 0 (0, 1) | 0 (0, 1) | 0 (0, 1) | 6 (4, 8) | 0 (0, 0) | 0 (0, 0) | 0 (0, 0) | 3 (1, 5) | 38 (33, 43) |
| 13 | 0 (0, 1) | 0 (0, 0) | 23 (8, 44) | 48 (27, 69) | 0 (0, 0) | 0 (0, 0) | 0 (0, 0) | 28 (12, 50) | 33 (15, 55) |
| 14 | 0 (0, 1) | 78 (51, 95) | 46 (20, 73) | 78 (51, 95) | 0 (0, 0) | 78 (51, 95) | 30 (9, 58) | 54 (27, 80) | 46 (20, 73) |
| 21 | 0 (0, 1) | 4 (0, 18) | 0 (0, 0) | 0 (0, 0) | 0 (0, 0) | 0 (0, 0) | 0 (0, 0) | 0 (0, 0) | 14 (3, 34) |
| 28A | 0 (0, 1) | 0 (0, 0) | 20 (8, 37) | 13 (4, 28) | 0 (0, 0) | 0 (0, 0) | 0 (0, 0) | 20 (8, 37) | 20 (8, 37) |
| 35D | 0 (0, 1) | 89 (67, 98) | 0 (0, 0) | 11 (2, 33) | 0 (0, 0) | 89 (67, 98) | 89 (67, 98) | 0 (0, 0) | 76 (51, 93) |
| 37 | 0 (0, 1) | 0 (0, 0) | 0 (0, 0) | 0 (0, 0) | 0 (0, 0) | 0 (0, 0) | 0 (0, 0) | 0 (0, 0) | 0 (0, 0) |
| 41A | 0 (0, 1) | 10 (10, 11) [c] | 9 (9, 10) [c] | 18 (17, 18) [c] | 0 (0, 0) [c] | 7 (7, 8) [c] | 8 (7, 8) [c] | 12 (11, 12) [c] | 30 (29, 30) [c] |
| 6H | 0 (0, 1) | 10 (10, 11) [c] | 9 (9, 10) [c] | 18 (17, 18) [c] | 0 (0, 0) [c] | 7 (7, 8) [c] | 8 (7, 8) [c] | 12 (11, 12) [c] | 30 (29, 30) [c] |
| NT | 0 (0, 1) | 10 (0, 49) | 26 (3, 67) | 10 (0, 49) | 0 (0, 0) | 10 (0, 49) | 10 (0, 49) | 42 (10, 80) | 42 (10, 80) |
| 10B | 0 (0, 0) | 10 (10, 11) [c] | 9 (9, 10) [c] | 18 (17, 18) [c] | 0 (0, 0) [c] | 7 (7, 8) [c] | 8 (7, 8) [c] | 12 (11, 12) [c] | 30 (29, 30) [c] |
| 10C | 0 (0, 0) | 10 (10, 11) [c] | 9 (9, 10) [c] | 18 (17, 18) [c] | 0 (0, 0) [c] | 7 (7, 8) [c] | 8 (7, 8) [c] | 12 (11, 12) [c] | 30 (29, 30) [c] |
| 10F | 0 (0, 0) | 0 (0, 0) | 0 (0, 0) | 20 (3, 55) | 0 (0, 0) | 0 (0, 0) | 0 (0, 0) | 0 (0, 0) | 0 (0, 0) |
| 11B | 0 (0, 0) | 10 (10, 11) [c] | 9 (9, 10) [c] | 18 (17, 18) [c] | 0 (0, 0) [c] | 7 (7, 8) [c] | 8 (7, 8) [c] | 12 (11, 12) [c] | 30 (29, 30) [c] |
| 11C | 0 (0, 0) | 10 (10, 11) [c] | 9 (9, 10) [c] | 18 (17, 18) [c] | 0 (0, 0) [c] | 7 (7, 8) [c] | 8 (7, 8) [c] | 12 (11, 12) [c] | 30 (29, 30) [c] |
| 11D | 0 (0, 0) | 10 (10, 11) [c] | 9 (9, 10) [c] | 18 (17, 18) [c] | 0 (0, 0) [c] | 7 (7, 8) [c] | 8 (7, 8) [c] | 12 (11, 12) [c] | 30 (29, 30) [c] |
| 11E | 0 (0, 0) | 10 (10, 11) [c] | 9 (9, 10) [c] | 18 (17, 18) [c] | 0 (0, 0) [c] | 7 (7, 8) [c] | 8 (7, 8) [c] | 12 (11, 12) [c] | 30 (29, 30) [c] |
| 11F | 0 (0, 0) | 10 (10, 11) [c] | 9 (9, 10) [c] | 18 (17, 18) [c] | 0 (0, 0) [c] | 7 (7, 8) [c] | 8 (7, 8) [c] | 12 (11, 12) [c] | 30 (29, 30) [c] |
| 12A | 0 (0, 0) | 10 (10, 11) [c] | 9 (9, 10) [c] | 18 (17, 18) [c] | 0 (0, 0) [c] | 7 (7, 8) [c] | 8 (7, 8) [c] | 12 (11, 12) [c] | 30 (29, 30) [c] |
| 12B | 0 (0, 0) | 10 (10, 11) [c] | 9 (9, 10) [c] | 18 (17, 18) [c] | 0 (0, 0) [c] | 7 (7, 8) [c] | 8 (7, 8) [c] | 12 (11, 12) [c] | 30 (29, 30) [c] |

| Serotype | % of pneumococcal cases (95% CI)[a] | % of isolates nonsusceptible to antibiotic classes, all adults (95% CI)[b] | | | | | | | |
|---|---|---|---|---|---|---|---|---|---|
| | | *Cephalosporin* | *Lincosamide* | *Antifolate* | *Fluoroquino-lone* | *Carbapenem* | *Penicillin* | *Tetracycline* | *Macrolide* |
| 15D | 0 (0, 0) | 0 (0, 0) | 0 (0, 0) | 0 (0, 0) | 0 (0, 0) | 0 (0, 0) | 0 (0, 0) | 0 (0, 0) | 100 (100, 100) |
| 15F | 0 (0, 0) | 0 (0, 0) | 100 (100, 100) | 0 (0, 0) | 0 (0, 0) | 0 (0, 0) | 0 (0, 0) | 0 (0, 0) | 100 (100, 100) |
| 16A | 0 (0, 0) | 10 (10, 11) [c] | 9 (9, 10) [c] | 18 (17, 18) [c] | 0 (0, 0) [c] | 7 (7, 8) [c] | 8 (7, 8) [c] | 12 (11, 12) [c] | 30 (29, 30) [c] |
| 17A | 0 (0, 0) | 10 (10, 11) [c] | 9 (9, 10) [c] | 18 (17, 18) [c] | 0 (0, 0) [c] | 7 (7, 8) [c] | 8 (7, 8) [c] | 12 (11, 12) [c] | 30 (29, 30) [c] |
| 18A | 0 (0, 0) | 0 (0, 0) | 0 (0, 0) | 0 (0, 0) | 0 (0, 0) | 0 (0, 0) | 0 (0, 0) | 0 (0, 0) | 0 (0, 0) |
| 18B | 0 (0, 0) | 0 (0, 0) | 0 (0, 0) | 50 (0, 100) | 0 (0, 0) | 0 (0, 0) | 0 (0, 0) | 0 (0, 0) | 0 (0, 0) |
| 18C | 0 (0, 0) | 0 (0, 0) | 15 (0, 65) | 85 (35, 100) | 0 (0, 0) | 0 (0, 0) | 0 (0, 0) | 85 (35, 100) | 85 (35, 100) |
| 18F | 0 (0, 0) | 10 (10, 11) [c] | 9 (9, 10) [c] | 18 (17, 18) [c] | 0 (0, 0) [c] | 7 (7, 8) [c] | 8 (7, 8) [c] | 12 (11, 12) [c] | 30 (29, 30) [c] |
| 19B | 0 (0, 0) | 10 (10, 11) [c] | 9 (9, 10) [c] | 18 (17, 18) [c] | 0 (0, 0) [c] | 7 (7, 8) [c] | 8 (7, 8) [c] | 12 (11, 12) [c] | 30 (29, 30) [c] |
| 19C | 0 (0, 0) | 10 (10, 11) [c] | 9 (9, 10) [c] | 18 (17, 18) [c] | 0 (0, 0) [c] | 7 (7, 8) [c] | 8 (7, 8) [c] | 12 (11, 12) [c] | 30 (29, 30) [c] |
| 2 | 0 (0, 0) | 10 (10, 11) [c] | 9 (9, 10) [c] | 18 (17, 18) [c] | 0 (0, 0) [c] | 7 (7, 8) [c] | 8 (7, 8) [c] | 12 (11, 12) [c] | 30 (29, 30) [c] |
| 22A | 0 (0, 0) | 10 (10, 11) [c] | 9 (9, 10) [c] | 18 (17, 18) [c] | 0 (0, 0) [c] | 7 (7, 8) [c] | 8 (7, 8) [c] | 12 (11, 12) [c] | 30 (29, 30) [c] |
| 23F | 0 (0, 0) | 31 (4, 75) | 31 (4, 75) | 69 (25, 96) | 0 (0, 0) | 31 (4, 75) | 0 (0, 0) | 50 (12, 88) | 31 (4, 75) |
| 24A | 0 (0, 0) | 0 (0, 0) | 0 (0, 0) | 0 (0, 0) | 0 (0, 0) | 0 (0, 0) | 0 (0, 0) | 0 (0, 0) | 0 (0, 0) |
| 24B | 0 (0, 0) | 10 (10, 11) [c] | 9 (9, 10) [c] | 18 (17, 18) [c] | 0 (0, 0) [c] | 7 (7, 8) [c] | 8 (7, 8) [c] | 12 (11, 12) [c] | 30 (29, 30) [c] |
| 24C | 0 (0, 0) | 10 (10, 11) [c] | 9 (9, 10) [c] | 18 (17, 18) [c] | 0 (0, 0) [c] | 7 (7, 8) [c] | 8 (7, 8) [c] | 12 (11, 12) [c] | 30 (29, 30) [c] |
| 25A | 0 (0, 0) | 10 (10, 11) [c] | 9 (9, 10) [c] | 18 (17, 18) [c] | 0 (0, 0) [c] | 7 (7, 8) [c] | 8 (7, 8) [c] | 12 (11, 12) [c] | 30 (29, 30) [c] |
| 25F | 0 (0, 0) | 10 (10, 11) [c] | 9 (9, 10) [c] | 18 (17, 18) [c] | 0 (0, 0) [c] | 7 (7, 8) [c] | 8 (7, 8) [c] | 12 (11, 12) [c] | 30 (29, 30) [c] |
| 27 | 0 (0, 0) | 10 (10, 11) [c] | 9 (9, 10) [c] | 18 (17, 18) [c] | 0 (0, 0) [c] | 7 (7, 8) [c] | 8 (7, 8) [c] | 12 (11, 12) [c] | 30 (29, 30) [c] |
| 28F | 0 (0, 0) | 10 (10, 11) [c] | 9 (9, 10) [c] | 18 (17, 18) [c] | 0 (0, 0) [c] | 7 (7, 8) [c] | 8 (7, 8) [c] | 12 (11, 12) [c] | 30 (29, 30) [c] |
| 29 | 0 (0, 0) | 10 (10, 11) [c] | 9 (9, 10) [c] | 18 (17, 18) [c] | 0 (0, 0) [c] | 7 (7, 8) [c] | 8 (7, 8) [c] | 12 (11, 12) [c] | 30 (29, 30) [c] |
| 32A | 0 (0, 0) | 10 (10, 11) [c] | 9 (9, 10) [c] | 18 (17, 18) [c] | 0 (0, 0) [c] | 7 (7, 8) [c] | 8 (7, 8) [c] | 12 (11, 12) [c] | 30 (29, 30) [c] |
| 32F | 0 (0, 0) | 10 (10, 11) [c] | 9 (9, 10) [c] | 18 (17, 18) [c] | 0 (0, 0) [c] | 7 (7, 8) [c] | 8 (7, 8) [c] | 12 (11, 12) [c] | 30 (29, 30) [c] |
| 33A | 0 (0, 0) | 0 (0, 0) | 0 (0, 0) | 0 (0, 0) | 0 (0, 0) | 0 (0, 0) | 0 (0, 0) | 0 (0, 0) | 0 (0, 0) |
| 33B | 0 (0, 0) | 10 (10, 11) [c] | 9 (9, 10) [c] | 18 (17, 18) [c] | 0 (0, 0) [c] | 7 (7, 8) [c] | 8 (7, 8) [c] | 12 (11, 12) [c] | 30 (29, 30) [c] |
| 33C | 0 (0, 0) | 10 (10, 11) [c] | 9 (9, 10) [c] | 18 (17, 18) [c] | 0 (0, 0) [c] | 7 (7, 8) [c] | 8 (7, 8) [c] | 12 (11, 12) [c] | 30 (29, 30) [c] |
| 33D | 0 (0, 0) | 10 (10, 11) [c] | 9 (9, 10) [c] | 18 (17, 18) [c] | 0 (0, 0) [c] | 7 (7, 8) [c] | 8 (7, 8) [c] | 12 (11, 12) [c] | 30 (29, 30) [c] |

| Serotype | % of pneumococcal cases (95% CI)[a] | % of isolates nonsusceptible to antibiotic classes, all adults (95% CI)[b] | | | | | | | |
|---|---|---|---|---|---|---|---|---|---|
| | | *Cephalosporin* | *Lincosamide* | *Antifolate* | *Fluoroquino-lone* | *Carbapenem* | *Penicillin* | *Tetracycline* | *Macrolide* |
| 35A | 0 (0, 0) | 0 (0, 0) | 0 (0, 0) | 100 (100, 100) | 0 (0, 0) | 0 (0, 0) | 0 (0, 0) | 100 (100, 100) | 0 (0, 0) |
| 35C | 0 (0, 0) | 10 (10, 11) [c] | 9 (9, 10) [c] | 18 (17, 18) [c] | 0 (0, 0) [c] | 7 (7, 8) [c] | 8 (7, 8) [c] | 12 (11, 12) [c] | 30 (29, 30) [c] |
| 36 | 0 (0, 0) | 10 (10, 11) [c] | 9 (9, 10) [c] | 18 (17, 18) [c] | 0 (0, 0) [c] | 7 (7, 8) [c] | 8 (7, 8) [c] | 12 (11, 12) [c] | 30 (29, 30) [c] |
| 39 | 0 (0, 0) | 10 (10, 11) [c] | 9 (9, 10) [c] | 18 (17, 18) [c] | 0 (0, 0) [c] | 7 (7, 8) [c] | 8 (7, 8) [c] | 12 (11, 12) [c] | 30 (29, 30) [c] |
| 40 | 0 (0, 0) | 10 (10, 11) [c] | 9 (9, 10) [c] | 18 (17, 18) [c] | 0 (0, 0) [c] | 7 (7, 8) [c] | 8 (7, 8) [c] | 12 (11, 12) [c] | 30 (29, 30) [c] |
| 41F | 0 (0, 0) | 10 (10, 11) [c] | 9 (9, 10) [c] | 18 (17, 18) [c] | 0 (0, 0) [c] | 7 (7, 8) [c] | 8 (7, 8) [c] | 12 (11, 12) [c] | 30 (29, 30) [c] |
| 42 | 0 (0, 0) | 10 (10, 11) [c] | 9 (9, 10) [c] | 18 (17, 18) [c] | 0 (0, 0) [c] | 7 (7, 8) [c] | 8 (7, 8) [c] | 12 (11, 12) [c] | 30 (29, 30) [c] |
| 43 | 0 (0, 0) | 10 (10, 11) [c] | 9 (9, 10) [c] | 18 (17, 18) [c] | 0 (0, 0) [c] | 7 (7, 8) [c] | 8 (7, 8) [c] | 12 (11, 12) [c] | 30 (29, 30) [c] |
| 44 | 0 (0, 0) | 10 (10, 11) [c] | 9 (9, 10) [c] | 18 (17, 18) [c] | 0 (0, 0) [c] | 7 (7, 8) [c] | 8 (7, 8) [c] | 12 (11, 12) [c] | 30 (29, 30) [c] |
| 45 | 0 (0, 0) | 100 (100, 100) | 0 (0, 0) | 100 (100, 100) | 0 (0, 0) | 0 (0, 0) | 0 (0, 0) | 0 (0, 0) | 0 (0, 0) |
| 46 | 0 (0, 0) | 10 (10, 11) [c] | 9 (9, 10) [c] | 18 (17, 18) [c] | 0 (0, 0) [c] | 7 (7, 8) [c] | 8 (7, 8) [c] | 12 (11, 12) [c] | 30 (29, 30) [c] |
| 47A | 0 (0, 0) | 10 (10, 11) [c] | 9 (9, 10) [c] | 18 (17, 18) [c] | 0 (0, 0) [c] | 7 (7, 8) [c] | 8 (7, 8) [c] | 12 (11, 12) [c] | 30 (29, 30) [c] |
| 47F | 0 (0, 0) | 10 (10, 11) [c] | 9 (9, 10) [c] | 18 (17, 18) [c] | 0 (0, 0) [c] | 7 (7, 8) [c] | 8 (7, 8) [c] | 12 (11, 12) [c] | 30 (29, 30) [c] |
| 48 | 0 (0, 0) | 10 (10, 11) [c] | 9 (9, 10) [c] | 18 (17, 18) [c] | 0 (0, 0) [c] | 7 (7, 8) [c] | 8 (7, 8) [c] | 12 (11, 12) [c] | 30 (29, 30) [c] |
| 6C | 0 (0, 0) | 30 (25, 36) | 7 (4, 10) | 27 (22, 32) | 0 (0, 0) | 7 (5, 10) | 3 (2, 6) | 4 (2, 7) | 48 (43, 54) |
| 6D | 0 (0, 0) | 50 (0, 100) | 50 (0, 100) | 50 (0, 100) | 0 (0, 0) | 0 (0, 0) | 0 (0, 0) | 0 (0, 0) | 100 (100, 100) |
| 7A | 0 (0, 0) | 10 (10, 11) [c] | 9 (9, 10) [c] | 18 (17, 18) [c] | 0 (0, 0) [c] | 7 (7, 8) [c] | 8 (7, 8) [c] | 12 (11, 12) [c] | 30 (29, 30) [c] |
| 7B | 0 (0, 0) | 0 (0, 0) | 0 (0, 0) | 100 (100, 100) | 0 (0, 0) | 0 (0, 0) | 0 (0, 0) | 100 (100, 100) | 0 (0, 0) |
| 7D | 0 (0, 0) | 10 (10, 11) [c] | 9 (9, 10) [c] | 18 (17, 18) [c] | 0 (0, 0) [c] | 7 (7, 8) [c] | 8 (7, 8) [c] | 12 (11, 12) [c] | 30 (29, 30) [c] |
| 9A | 0 (0, 0) | 10 (10, 11) [c] | 9 (9, 10) [c] | 18 (17, 18) [c] | 0 (0, 0) [c] | 7 (7, 8) [c] | 8 (7, 8) [c] | 12 (11, 12) [c] | 30 (29, 30) [c] |
| 9L | 0 (0, 0) | 27 (0, 91) | 0 (0, 0) | 27 (0, 91) | 0 (0, 0) | 0 (0, 0) | 0 (0, 0) | 27 (0, 91) | 0 (0, 0) |

Abbreviations: CI – confidence interval.
[a] Estimated using data from previously published data on pneumococcal pneumonia[2] and Active Bacterial Core surveillance data.
[b] Nonsusceptibility to antibiotic classes evaluated based on nonsusceptibility to specific agents within each class, as defined in **Table S2**.
[c] Imputed as the inverse variance-weighted average proportion of nonsusceptible isolates across all serotypes with known susceptibility.

**Table S5. Percent and burdens of antibiotic-nonsusceptible non-bacteremic pneumococcal pneumonia by nonsusceptibility criteria and patient age group**

| Nonsusceptible to | Age group | Percent of cases (95% CI)[a] | Outpatient visits | | Hospitalizations | |
|---|---|---|---|---|---|---|
| | | | *Incidence per 10,000 person-years (95% CI)* | *Annual no. in thousands (95% CI)* | *Incidence per 10,000 person-years (95% CI)* | *Annual no. in thousands (95% CI)* |
| ≥1 outpatient first-line antibiotic agent[b] | | | | | | |
| | 18-49 years | 35.2 (29.0, 42.1) | 4.9 (3.0, 7.6) | 67.8 (42.2, 104.6) | 0.1 (0.1, 0.2) | 1.6 (1.0, 2.4) |
| | 50-64 years | 30.0 (25.8, 34.7) | 21.1 (15.2, 28.9) | 132.5 (95.7, 182.0) | 0.8 (0.6, 1.0) | 4.9 (3.7, 6.3) |
| | ≥65 years | 30.9 (25.6, 36.8) | 47.9 (35.2, 64.7) | 258.8 (190.4, 349.9) | 2.4 (1.8, 3.2) | 13.2 (9.9, 17.1) |
| ≥2 outpatient first-line antibiotic agents[b] | | | | | | |
| | 18-49 years | 17.9 (13.5, 23.8) | 2.5 (1.5, 4.1) | 34.6 (20.6, 56.3) | 0.1 (0.1, 0.1) | 0.8 (0.5, 1.3) |
| | 50-64 years | 14.4 (11.2, 18.6) | 10.1 (6.9, 14.8) | 63.7 (43.5, 93.1) | 0.4 (0.3, 0.5) | 2.3 (1.7, 3.3) |
| | ≥65 years | 12.4 (9.3, 16.7) | 19.2 (13.2, 28.2) | 103.9 (71.2, 152.3) | 1.0 (0.7, 1.4) | 5.3 (3.7, 7.5) |
| ≥1 inpatent first-line antibiotic agent[c] | | | | | | |
| | 18-49 years | 18.1 (12.5, 25.5) | 2.5 (1.4, 4.3) | 34.8 (19.8, 58.9) | 0.1 (0.0, 0.1) | 0.8 (0.5, 1.4) |
| | 50-64 years | 11.1 (8.0, 15.3) | 7.8 (5.0, 11.9) | 48.9 (31.7, 75.0) | 0.3 (0.2, 0.4) | 1.8 (1.2, 2.6) |
| | ≥65 years | 13.6 (9.5, 19.2) | 21.1 (13.7, 32.3) | 114.0 (74.2, 173.8) | 1.1 (0.7, 1.6) | 5.8 (3.8, 8.6) |
| >1 antibiotic class | | | | | | |
| | 18-49 years | 27.1 (21.3, 34.6) | 3.8 (2.3, 6.0) | 52.4 (31.8, 83.1) | 0.1 (0.1, 0.1) | 1.2 (0.7, 1.9) |
| | 50-64 years | 20.4 (16.5, 25.7) | 14.4 (10.0, 20.6) | 90.3 (62.8, 129.7) | 0.5 (0.4, 0.7) | 3.3 (2.4, 4.5) |
| | ≥65 years | 20.9 (16.0, 27.2) | 32.5 (22.6, 46.4) | 175.6 (122.4, 250.7) | 1.7 (1.2, 2.3) | 8.9 (6.4, 12.4) |
| ≥3 antibiotic classes | | | | | | |
| | 18-49 years | 22.1 (16.6, 29.5) | 3.1 (1.8, 5.0) | 42.8 (25.4, 69.7) | 0.1 (0.0, 0.1) | 1.0 (0.6, 1.6) |
| | 50-64 years | 16.4 (12.7, 21.3) | 11.5 (7.8, 16.9) | 72.5 (49.3, 106.4) | 0.4 (0.3, 0.6) | 2.7 (1.9, 3.7) |
| | ≥65 years | 15.1 (11.2, 20.4) | 23.4 (15.9, 34.5) | 126.5 (86.1, 186.3) | 1.2 (0.8, 1.7) | 6.4 (4.5, 9.2) |

Abbreviations: CI – confidence interval; No. – number
[a] Estimated as annual no. nonsusceptible pneumococcal outpatient visits for each nonsusceptibility category divided by annual no. total pneumococcal-attributable outpatient visits for each age group.
[b] Outpatient first-line antibiotic agents for adult pneumonia defined as amoxicillin, doxycycline, and erythromycin (proxy for all macrolides).[5]
[c] Inpatient first-line antibiotic agents for adult pneumonia defined as cephalosporins (cefotaxime, ceftriaxone, and cefuroxime) and levofloxacin (proxy for all respiratory fluoroquinolones).[5]

**Table S6. Percent and burdens of antibiotic-nonsusceptible non-bacteremic pneumococcal pneumonia by drug class and patient age group**

| | | | **Outpatient visits** | | **Hospitalizations** | |
|---|---|---|---|---|---|---|
| **Nonsusceptible to[a]** | **Age group** | **Percent of cases (95% CI)[b]** | *Incidence per 10,000 person-years (95% CI)* | *Annual no. in thousands (95% CI)* | *Incidence per 10,000 person-years (95% CI)* | *Annual no. in thousands (95% CI)* |
| Macrolides | | | | | | |
| | 18-49 years | 32.0 (26.3, 38.2) | 4.5 (2.8, 6.9) | 61.7 (38.3, 95.2) | 0.1 (0.1, 0.2) | 1.5 (0.9, 2.2) |
| | 50-64 years | 27.3 (23.3, 31.7) | 19.2 (13.5, 26.4) | 120.7 (86.9, 166.2) | 0.7 (0.5, 0.9) | 4.5 (3.4, 5.8) |
| | ≥65 years | 27.4 (22.5, 32.7) | 42.5 (31.2, 57.5) | 229.8 (168.6, 310.7) | 2.2 (1.6, 2.8) | 11.7 (8.8, 15.2) |
| Antifolates | | | | | | |
| | 18-49 years | 23.9 (18.9, 29.6) | 3.3 (2.0, 5.2) | 46.2 (28.2, 72.5) | 0.1 (0.0, 0.1) | 1.1 (0.7, 1.7) |
| | 50-64 years | 17.9 (14.3, 22.3) | 12.6 (8.7, 18.1) | 79.3 (54.9, 113.6) | 0.5 (0.6, 0.6) | 2.9 (2.1, 4.0) |
| | ≥65 years | 21.9 (17.4, 27.5) | 34.1 (24.4, 47.5) | 184.0 (131.7, 256.7) | 1.7 (1.3, 2.3) | 9.4 (6.9, 12.6) |
| Cephalosporins | | | | | | |
| | 18-49 years | 18.1 (12.4, 25.5) | 2.5 (1.4, 4.3) | 34.8 (19.8, 58.8) | 0.1 (0.0, 0.1) | 0.8 (0.5, 1.4) |
| | 50-64 years | 11.0 (8.0, 15.3) | 7.8 (5.0, 11.9) | 48.8 (31.6, 74.8) | 0.3 (0.2, 0.4) | 1.8 (1.2, 2.6) |
| | ≥65 years | 13.5 (9.4, 19.1) | 21.0 (13.6, 32.0) | 113.3 (73.6 173.0) | 1.1 (0.7, 1.6) | 5.8 (3.8, 8.6) |
| Tetracyclines | | | | | | |
| | 18-49 years | 14.1 (10.7, 18.3) | 2.0 (1.2, 3.2) | 27.1 (16.3, 43.6) | 0.0 (0.0, 0.1) | 0.6 (0.4, 1.0) |
| | 50-64 years | 13.5 (10.8, 17.0) | 9.5 (6.6, 13.6) | 59.6 (41.4, 85.9) | 0.3 (0.3, 0.5) | 2.2 (1.6, 3.0) |
| | ≥65 years | 12.0 (9.0, 16.7) | 18.8 (12.8, 28.1) | 101.4 (69.3, 151.7) | 1.0 (0.7, 1.4) | 5.2 (3.6, 7.5) |
| Carbepenems | | | | | | |
| | 18-49 years | 15.3 (9.8, 22.8) | 2.1 (1.2, 3.7) | 29.4 (16.0, 51.7) | 0.1 (0.0, 0.1) | 0.7 (0.4, 1.2) |
| | 50-64 years | 8.2 (5.3, 12.4) | 5.7 (3.4, 9.4) | 36.0 (21.7, 59.4) | 0.2 (0.1, 0.3) | 1.3 (0.8, 2.1) |

| Nonsusceptible to[a] | Age group | Percent of cases (95% CI)[b] | Outpatient visits | | Hospitalizations | |
|---|---|---|---|---|---|---|
| | | | *Incidence per 10,000 person-years (95% CI)* | *Annual no. in thousands (95% CI)* | *Incidence per 10,000 person-years (95% CI)* | *Annual no. in thousands (95% CI)* |
| | ≥65 years | 9.5 (6.1, 14.6) | 14.8 (8.9, 24.1) | 79.7 (47.9, 130.4) | 0.7 (0.5, 1.2) | 4.1 (2.5, 6.5) |
| Penicillins | | | | | | |
| | 18-49 years | 14.7 (10.0, 21.5) | 2.1 (1.2, 3.6) | 28.4 (16.0, 49.1) | 0.1 (0.0, 0.1) | 0.7 (0.4, 1.1) |
| | 50-64 years | 7.9 (5.3, 11.7) | 5.5 (3.4, 9.0) | 34.7 (21.3 56.5) | 0.2 (0.1, 0.3) | 1.3 (0.8, 2.0) |
| | ≥65 years | 7.6 (4.7, 12.0) | 11.8 (6.9, 19.6) | 63.7 (37.5, 106.1) | 0.6 (0.4, 1.0) | 3.2 (1.9, 5.3) |
| Lincosamides | | | | | | |
| | 18-49 years | 10.2 (7.3, 14.5) | 1.4 (0.8, 2.4) | 19.7 (11.4, 33.3) | 0.0 (0.0, 0.1) | 0.5 (0.3, 0.8) |
| | 50-64 years | 10.3 (7.9, 14.0) | 7.3 (4.9, 11.0) | 45.8 (30.7, 69.2) | 0.3 (0.2, 0.4) | 1.7 (1.2, 2.4) |
| | ≥65 years | 7.2 (5.2, 10.7) | 11.3 (7.5, 17.7) | 60.9 (40.3 95.8) | 0.6 (0.4, 0.9) | 3.1 (2.1, 4.8) |
| Fluoroquinolones | | | | | | |
| | 18-49 years | 0.2 (0.1, 0.5) | 0.0 (0.0, 0.1) | 0.4 (0.1, 1.1) | 0.0 (0.0, 0.0) | 0.0 (0.0, 0.0) |
| | 50-64 years | 0.2 (0.1, 0.4) | 0.1 (0.0, 0.3) | 0.7 (0.3, 1.7) | 0.0 (0.0, 0.0) | 0.0 (0.0, 0.1) |
| | ≥65 years | 0.1 (0.0, 0.2) | 0.1 (0.0, 0.4) | 0.6 (0.2, 1.9) | 0.0 (0.0, 0.0) | 0.0 (0.0, 0.1) |

Abbreviations: CI – confidence interval; No. – number

[a] Nonsusceptibility to each class based on nonsusceptibility to specific agents within each class, as defined in **Table S2**. Linezolids and glycopeptides not shown as all Active Bacterial Core surveillance isolates were susceptible to those classes.

[b] Estimated as annual no. nonsusceptible pneumococcal outpatient visits for each nonsusceptibility category divided by annual no. pneumococcal-attributable outpatient visits for each age group.

**Table S7. Percent and burdens of antibiotic-nonsusceptible pneumococcal sinusitis by nonsusceptibility criteria and patient age group**

| Nonsusceptible to | Age group | Percent of cases (95% CI)[a] | Outpatient visit incidence per 10,000 person-years (95% CI) | Annual no. outpatient visits in thousands (95% CI) |
|---|---|---|---|---|
| First-line antibiotic agent[b] | | | | |
| | 18-49 years | 11.0 (6.4, 17.8) | 38.9 (19.1, 72.6) | 538.0 (264.2, 1,004.3) |
| | 50-64 years | 5.9 (3.4, 9.7) | 24.0 (11.5, 46.0) | 151.2 (72.5, 289.0) |
| | ≥65 years | 6.7 (3.9, 11.0) | 26.1 (12.5, 50.3) | 140.9 (67.6, 271.6) |
| ≥1 alternative antibiotic agent[c] | | | | |
| | 18-49 years | 10.3 (7.4, 14.5) | 36.7 (20.2, 61.9) | 507.4 (280.0, 855.4) |
| | 50-64 years | 10.2 (7.7, 13.9) | 42.1 (23.5, 70.0) | 264.8 (147.9, 440.4) |
| | ≥65 years | 7.7 (5.5, 11.2) | 30.3 (16.4, 53.0) | 163.7 (88.6, 286.6) |
| >1 antibiotic class | | | | |
| | 18-49 years | 27.1 (21.3, 34.6) | 96.6 (55.6, 152.9) | 1,336.5 (769.1, 2,114.2) |
| | 50-64 years | 20.4 (16.5, 25.7) | 83.8 (48.1, 133.3) | 526.8 (302.5, 838.5) |
| | ≥65 years | 20.9 (16.0, 27.2) | 82.4 (46.2, 135.0) | 445.2 (249.4, 729.4) |
| ≥3 antibiotic classes | | | | |
| | 18-49 years | 22.1 (16.6, 29.5) | 78.9 (44.5, 128.4) | 1,091.2 (615.3, 1,775.8) |
| | 50-64 years | 16.4 (12.7, 21.3) | 67.2 (38.1, 109.0) | 422.9 (239.7, 685.6) |
| | ≥65 years | 15.1 (11.2, 20.4) | 59.3 (32.8, 99.5) | 320.7 (177.1, 537.7) |

Abbreviations: CI – confidence interval; No. – number.

[a] Estimated as annual no. nonsusceptible pneumococcal outpatient visits for each nonsusceptibility category divided by annual no. total pneumococcal-attributable outpatient visits for each age group.

[b] First-line antibiotic agent for adult sinusitis defined as amoxicillin.[6,7]

[c] Alternative antibiotic agents for adult sinusitis defined as doxycycline, levofloxacin, clindamycin, and cephalosporins.[6,7]

**Table S8. Percent and burdens of antibiotic-nonsusceptible pneumococcal sinusitis by drug class and patient age group**

| Nonsusceptible to[a] | Age group | Percent of cases (95% CI)[b] | Outpatient visit incidence per 10,000 person-years (95% CI) | Annual no. outpatient visits in thousands (95% CI) |
|---|---|---|---|---|
| Macrolides | | | | |
| | 18-49 years | 32.0 (26.3, 38.2) | 114.1 (66.9, 174.9) | 1,577.4 (925.2, 2,418.6) |
| | 50-64 years | 27.3 (23.3, 31.7) | 112.1 (65.6, 172.5) | 704.8 (412.9, 1,085.0) |
| | ≥65 years | 27.4 (22.5, 32.7) | 107.9 (62.2, 169.9) | 583.2 (336.0, 918.3) |
| Antifolatess | | | | |
| | 18-49 years | 23.9 (18.9, 29.6) | 85.3 (49.4, 133.4) | 1,180.3 (682.8, 1,845.3) |
| | 50-64 years | 17.9 (14.3, 22.3) | 73.5 (42.1, 116.9) | 462.3 (264.8, 735.5) |
| | ≥65 years | 21.9 (17.4, 27.5) | 86.4 (49.1, 139.1) | 466.9 (265.4, 751.6) |
| Cephalosporins | | | | |
| | 18-49 years | 18.1 (12.4 25.5 | 64.2 (34.9, 108.4) | 888.0 (482.2, 1499.3) |
| | 50-64 years | 11.0 (8.0 15.3 | 45.2 (24.8, 76.0) | 284.0 (156.1, 478.2) |
| | ≥65 years | 13.5 (9.4 19.1 | 53.1 (28.5, 91.7) | 286.7 (153.8, 495.6) |
| Tetracyclines | | | | |
| | 18-49 years | 14.1 (10.7, 18.3) | 47.6 (26.3, 80.7) | 692.8 (394.4, 1111.0) |
| | 50-64 years | 13.5 (10.8, 17.0) | 55.3 (31.7, 88.3) | 347.9 (199.5, 555.2) |
| | ≥65 years | 12.0 (9.0, 16.7) | 50.1 (28.5, 80.3) | 257.3 (142.1, 436.0) |
| Carbepenems | | | | |
| | 18-49 years | 15.3 (9.8, 22.8) | 54.2 (28.3, 95.3) | 749.0 (391.1, 1318.1) |
| | 50-64 years | 8.2 (5.3, 12.4) | 33.3 (17.4, 59.7) | 209.5 (109.3, 375.4) |
| | ≥65 years | 9.5 (6.1, 14.6) | 37.3 (19.0, 68.2) | 201.5 (102.7, 368.5) |
| Penicillins | | | | |
| | 18-49 years | 14.7 (10.0, 21.5) | 52.3 (28.2, 90.5) | 723.7 (390.0, 1251.2) |
| | 50-64 years | 7.9 (5.3, 11.7) | 32.1 (17.0, 56.9) | 202.0 (106.7, 357.6) |
| | ≥65 years | 7.6 (4.7, 12.0) | 29.8 (15.0, 55.2) | 160.8 (80.9, 298.4) |
| Lincosamides | | | | |
| | 18-49 years | 10.2 (7.3, 14.5) | 36.3 (20.0, 61.4) | 502.2 (277.3, 849.2) |
| | 50-64 years | 10.3 (7.9, 14.0) | 42.5 (23.8, 70.5) | 267.2 (149.7, 443.5) |
| | ≥65 years | 7.2 (5.2, 10.7) | 28.6 (15.5, 50.3) | 154.4 (83.6, 271.6) |
| Fluoroquinolones | | | | |
| | 18-49 years | 0.2 (0.1, 0.5) | 0.7 (0.2, 2.0) | 9.9 (3.1, 27.3) |
| | 50-64 years | 0.2 (0.1, 0.4) | 0.6 (0.2, 1.6) | 4.0 (1.4, 10.2) |
| | ≥65 years | 0.1 (0.0, 0.2) | 0.3 (0.1, 0.9) | 1.6 (0.5, 5.1) |

Abbreviations: CI – confidence interval; No. – number

[a] Nonsusceptibility to each class based on nonsusceptibility to specific agents within each class, as defined in **Table S2**. Linezolids and glycopeptides not shown as all Active Bacterial Core surveillance isolates were susceptible to those classes.
[b] Estimated as annual no. nonsusceptible pneumococcal outpatient visits for each nonsusceptibility category divided by annual no. pneumococcal-attributable outpatient visits for each age group.

**Table S9. Burdens of antibiotic-nonsusceptible non-bacteremic pneumococcal pneumonia by antibiotic class and PCV-targeted serotypes**

| | | | Outpatient visits | | Hospitalizations | |
|---|---|---|---|---|---|---|
| **Nonsusceptible to**[a] | **PCV-targeted serotypes**[b] | **Percent of cases (95% CI)**[c] | *Incidence per 10,000 person-years (95% CI)* | *Annual no. in thousands (95% CI)* | *Incidence per 10,000 person-years (95% CI)* | *Annual no. in thousands (95% CI)* |
| Macrolides | | | | | | |
| | PCV15 | 14.0 (10.4 18.6) | 8.1 (5.8 11.4) | 207.4 (147.1, 289.8) | 3.5 (2.5 4.8) | 8.9 (6.3, 12.3) |
| | PCV20 | 17.1 (13.4 21.5) | 9.9 (7.4 13.3) | 253.8 (188.3, 338.3) | 4.3 (3.2 5.7) | 10.9 (8.2, 14.4) |
| | PCV21 | 22.9 (18.6 27.8) | 13.3 (10.2 17.2) | 338.8 (259.4, 439.1) | 5.7 (4.4 7.3) | 14.5 (11.2, 18.5) |
| | NVT | 1.5 (0.7 4.6) | 0.9 (0.4 2.7) | 22.2 (10.2, 67.9) | 0.4 (0.2 1.1) | 0.9 (0.4, 2.8) |
| Antifolates | | | | | | |
| | PCV15 | 13.1 (9.3, 18.2) | 7.6 (5.2, 11.0) | 194.6 (132.3, 281.8) | 0.3 (0.2, 0.5) | 8.6 (5.9, 12.3) |
| | PCV20 | 14.8 (11.0, 19.7) | 8.6 (6.1, 12.1) | 219.8 (155.9, 308.1) | 0.4 (0.3, 0.5) | 9.7 (6.9, 13.4) |
| | PCV21 | 10.8 (8.0, 14.5) | 6.3 (4.4, 8.8) | 159.9 (113.1, 225.3) | 0.3 (0.2, 0.4) | 6.7 (4.7, 9.4) |
| | NVT | 1.4 (0.6, 3.2) | 0.8 (0.4, 1.9) | 20.7 (9.4, 47.5) | 0.0 (0.0, 0.1) | 0.9 (0.4, 2.0) |
| Cephalosporins | | | | | | |
| | PCV15 | 8.0 (5.1, 11.8) | 4.6 (2.9, 7.1) | 118.0 (72.9, 181.5) | 0.2 (0.1, 0.3) | 5.01 (3.1, 7.8) |
| | PCV20 | 8.2 (5.4, 12.1) | 4.8 (3.0, 7.3) | 122.2 (77.2, 185.8) | 0.2 (0.1, 0.3) | 5.3 (3.3, 8.0) |
| | PCV21 | 10.7 (7.2, 15.8) | 6.2 (4.1, 9.5) | 159.1 (103.6, 242.8) | 0.3 (0.2, 0.4) | 6.8 (4.4, 10.3) |
| | NVT | 0.4 (0.2, 1.6) | 0.3 (0.1, 0.9) | 6.6 (2.4, 23.2) | 0.0 (0.0, 0.0) | 0.3 (0.1, 1.0) |
| Tetracyclines | | | | | | |
| | PCV15 | 7.5 (5.1, 11.1) | 4.4 (2.8, 6.7) | 111.2 (72.6, 171.0) | 0.2 (0.1, 0.3) | 4.8 (3.1, 7.4) |
| | PCV20 | 8.6 (6.1, 12.2) | 5.0 (3.4 7.4) | 127.5 (87.1, 188.2) | 0.2 (0.1, 0.3) | 5.5 (3.7, 8.1) |

| Nonsusceptible to[a] | PCV-targeted serotypes[b] | Percent of cases (95% CI)[c] | Outpatient visits | | Hospitalizations | |
|---|---|---|---|---|---|---|
| | | | *Incidence per 10,000 person-years (95% CI)* | *Annual no. in thousands (95% CI)* | *Incidence per 10,000 person-years (95% CI)* | *Annual no. in thousands (95% CI)* |
| Carbapenems | PCV21 | 8.6 (6.6, 11.4) | 5.0 (3.6 6.9) | 127.6 (92.7, 176.9) | 0.2 (0.2, 0.3) | 5.3 (3.9, 7.3) |
| | NVT | 0.9 (0.4, 2.2) | 0.5 (0.2 1.3) | 13.7 (6.3, 33.5) | 0.0 (0.0, 0.1) | 0.6 (0.3, 1.4) |
| Penicillins | PCV15 | 5.0 (2.8, 8.4) | 2.9 (1.6, 5.0) | 74.3 (40.1, 127.7) | 0.1 (0.1, 0.2) | 3.1 (1.7, 5.4) |
| | PCV20 | 5.3 (3.0, 8.6) | 3.1 (1.7, 5.2) | 78.3 (44.1, 131.5) | 0.1 (0.1, 0.2) | 3.3 (1.9, 5.6) |
| | PCV21 | 8.0 (4.8, 12.7) | 4.6 (2.7, 7.6) | 118.2 (69.5, 194.1) | 0.2 (0.1, 0.3) | 75.0 (2.9, 8.1) |
| | NVT | 0.2 (0.0, 0.8) | 0.1 (0.0, 0.5) | 2.6 (0.3, 12.5) | 0.0 (0.0, 0.0) | 0.1 (0.0, 0.5) |
| Lincosamides | PCV15 | 3.2 (1.5, 5.7) | 1.8 (0.9, 3.4) | 46.7 (22.5, 86.9) | 0.1 (0.0, 0.1) | 1.9 (0.9, 3.6) |
| | PCV20 | 3.4 (1.7, 5.9) | 1.9 (1.0, 3.5) | 49.7 (25.3, 89.8) | 0.1 (0.0, 0.1) | 2.0 (1.0, 3.7) |
| | PCV21 | 7.7 (4.8, 12.2) | 4.5 (2.7, 7.3) | 114.4 (69.0, 185.3) | 0.2 (0.1, 0.3) | 4.7 (2.8, 7.6) |
| | NVT | 0.2 (0.0, 0.8) | 0.1 (0.0, 0.5) | 2.6 (0.3, 12.5) | 0.0 (0.0, 0.0) | 0.1 (0.0, 0.5) |
| Fluoroquinolones | PCV15 | 5.1 (3.2, 8.0) | 2.9 (1.8 4.8) | 75.2 (45.9, 121.9) | 0.1 (0.1, 0.2) | 3.1 (1.9, 5.1) |
| | PCV20 | 5.5 (3.6 8.3) | 3.2 (2.0 5.0) | 80.9 (51.8, 127.3) | 0.1 (0.1, 0.2) | 3.4 (2.2, 5.3) |
| | PCV21 | 6.6 (4.8 9.3) | 3.8 (2.7 5.7) | 98.2 (68.2, 144.3) | 0.2 (0.1, 0.2) | 4.1 (2.9, 6.0) |
| | NVT | 0.6 (0.2 1.8) | 0.3 (0.1 1.0) | 8.5 (3.3, 26.6) | 0.0 (0.0, 0.0) | 0.3 (0.1, 1.1) |
| | PCV15 | 0.1 (0.0 0.2) | 0.0 (0.0 0.1) | 1.1 (0.4, 2.6) | 0.0 (0.0, 0.0) | 0.0 (0.0, 0.1) |
| | PCV20 | 0.1 (0.0 0.2) | 0.0 (0.0 0.1) | 1.1 (0.4, 2.6) | 0.0 (0.0, 0.0) | 0.0 (0.0, 0.1) |
| | PCV21 | 0.1 (0.1 0.2) | 0.1 (0.0 0.1) | 1.7 (0.7, 3.5) | 0.0 (0.0, 0.0) | 0.1 (0.0, 0.1) |
| | NVT | 0.0 (0.0, 0.0) | 0.0 (0.0, 0.0) | 0.0 (0.0, 0.2) | 0.0 (0.0, 0.0) | 0.0 (0.0, 0.0) |

Abbreviations: PCV – pneumococcal conjugate vaccine; CI – confidence interval; No. – number; NVT – non-vaccine type.

[a] Nonsusceptibility to each class based on nonsusceptibility to specific agents within each class, as defined in **Table S2**. Linezolids and glycopeptides not shown as all Active Bacterial Core surveillance isolates were susceptible to those classes.

[b] PCV15-targeted serotypes: 1, 3, 4, 5, 6A/C, 6B, 7F, 9V, 14, 18C, 19F, 19A, 22F, 23F, 33F. PCV20-targeted serotypes 1, 3, 4, 5, 6A/C, 6B, 7F, 8, 9V, 10A, 11A, 12F, 14, 15B, 18C, 19F, 19A, 22F, 23F, 33F. PCV21-targeted serotypes 3, 6A/C, 7F, 8, 9N, 10A, 11A, 12F, 15A, 15C, 16F, 17F, 19A, 20A, 22F, 23A, 23B, 24F, 31, 33F, 35B.

[c] Estimated as annual no. nonsusceptible pneumococcal outpatient visits for each nonsusceptibility category divided by annual no. total pneumococcal-attributable outpatient visits (N = 1,483,052 [95% CI 1,248,097, 1,760,694]).

**Table S10. Burdens of antibiotic-nonsusceptible pneumococcal sinusitis by antibiotic class and PCV-targeted serotypes**

| Nonsusceptible to[a] | PCV-targeted serotypes[b] | Percent of cases (95% CI)[c] | Outpatient visits | |
|---|---|---|---|---|
| | | | *Incidence per 10,000 person-years (95% CI)* | *Annual no. in thousands (95% CI)* |
| Macrolides | | | | |
| | PCV15 | 14.6 (10.4, 19.6) | 55.1 (31.1, 88.8) | 1,406.4 (793.3, 2,266.0) |
| | PCV20 | 17.2 (13.1, 22.0) | 65.0 (37.7, 101.5) | 1,659.8 (961.5, 2,589.6) |
| | PCV21 | 24.0 (19.4, 29.3) | 90.8 (53.7, 137.7) | 2,317.6 (1,371.5, 3,515.5) |
| | NVT | 1.7 (0.8, 5.2) | 6.4 (2.5, 20.1) | 163.8 (64.8, 512.7) |
| Antifolates | | | | |
| | PCV15 | 12.4 (8.6, 17.4) | 46.9 (25.8, 78.0) | 1,197.9 (659.2, 1,990.6) |
| | PCV20 | 13.9 (10.1, 18.8) | 52.5 (29.7, 85.0) | 1,340.4 (758.1, 2,168.6) |
| | PCV21 | 12.7 (9.5, 16.9) | 48.1 (27.5, 76.9) | 1,227.0 (701.7, 1,961.9) |
| | NVT | 1.6 (0.7, 3.8) | 5.9 (2.2, 15.1) | 149.5 (56.8, 385.1) |
| Cephalosporins | | | | |
| | PCV15 | 8.6 (5.4, 12.6) | 32.3 (16.9, 55.9) | 824.8 (431.2, 1,427.2) |
| | PCV20 | 8.8 (5.7, 12.8) | 33.1 (17.5, 56.9) | 845.6 (447.2, 1,451.9) |
| | PCV21 | 11.9 (7.9, 17.7) | 44.9 (24.2, 77.2) | 1,146.9 (617.4, 1,971.2) |
| | NVT | 0.4 (0.1, 1.7) | 1.6 (0.5, 6.5) | 41.0 (13.1, 165.6) |
| Tetracyclines | | | | |
| | PCV15 | 7.8 (5.3, 11.4) | 29.4 (16.0, 49.9) | 749.9 (407.9, 1,274.4) |
| | PCV20 | 8.7 (6.1, 12.2) | 32.9 (18.3, 54.5) | 840.5 (467.1, 1,391.0) |
| | PCV21 | 9.5 (7.1, 12.5) | 35.9 (20.6, 57.1) | 915.5 (524.8, 1,458.3) |
| | NVT | 1.0 (0.5, 2.6) | 3.9 (1.6, 10.4) | 99.9 (39.8, 266.1) |
| Carbapenems | | | | |
| | PCV15 | 6.0 (3.4, 9.9) | 22.7 (10.8, 42.7) | 578.2 (276.2, 1,091.1) |
| | PCV20 | 6.2 (3.5, 10.1) | 23.3 (11.4, 43.5) | 595.7 (290.2, 1,111.3) |
| | PCV21 | 9.4 (5.7, 15.0) | 35.4 (18.0, 64.2) | 903.6 (459.0, 1,639.8) |
| | NVT | 0.2 (0.0, 1.0) | 0.7 (0.1, 3.7) | 18.1 (2.1, 94.1) |
| Penicillins | | | | |
| | PCV15 | 4.0 (2.0, 6.9) | 14.9 (6.7, 29.5) | 380.4 (171.1, 753.9) |
| | PCV20 | 4.2 (2.3, 7.2) | 15.8 (7.4, 30.6) | 404.2 188.1, 782.3) |
| | PCV21 | 9.9 (6.3, 15.3) | 37.4 (19.7, 66.1) | 954.2 502.3, 1,686.9) |
| | NVT | 0.2 (0.0, 1.0) | 0.7 (0.1, 3.8) | 19.1 (2.2, 98.2) |
| Lincosamides | | | | |
| | PCV15 | 5.8 (3.6, 9.0) | 21.9 (11.4, 39.0) | 559.3 (291.3, 996.0) |

| Nonsusceptible to[a] | PCV-targeted serotypes[b] | Percent of cases (95% CI)[c] | Outpatient visits | |
|---|---|---|---|---|
| | | | *Incidence per 10,000 person-years (95% CI)* | *Annual no. in thousands (95% CI)* |
| | PCV20 | 6.1 (4.0, 9.3) | 23.2 (12.3, 40.4) | 590.9 (315.0, 1,032.3) |
| | PCV21 | 7.2 (5.1, 10.2) | 27.3 (15.2, 45.4) | 695.8 (387.8, 1,158.0) |
| | NVT | 0.7 (0.3, 2.1) | 2.6 (1.0, 8.7) | 67.1 (25.3, 221.5) |
| Fluoroquinolones | | | | |
| | PCV15 | 0.1 (0.0, 0.3) | 0.5 (0.1, 1.3) | 11.7 (3.7, 33.3) |
| | PCV20 | 0.1 (0.0, 0.3) | 0.5 (0.1, 1.3) | 11.7 (3.7, 33.3) |
| | PCV21 | 0.2 (0.1, 0.4) | 0.6 (0.2, 1.5) | 14.6 (5.3, 37.5) |
| | NVT | 0.0 (0.0, 0.0) | 0.0 (0.0, 0.0) | 0.2 (0.0, 1.2) |

Abbreviations: PCV – pneumococcal conjugate vaccine; CI – confidence interval; No. – number; NVT – non-vaccine type.

[a] Nonsusceptibility to each class based on nonsusceptibility to specific agents within each class, as defined in **Table S2**. Linezolids and glycopeptides not shown as all Active Bacterial Core surveillance isolates were susceptible to those classes.

[b] PCV15-targeted serotypes: 1, 3, 4, 5, 6A/C, 6B, 7F, 9V, 14, 18C, 19F, 19A, 22F, 23F, 33F. PCV20-targeted serotypes 1, 3, 4, 5, 6A/C, 6B, 7F, 8, 9V, 10A, 11A, 12F, 14, 15B, 18C, 19F, 19A, 22F, 23F, 33F. PCV21-targeted serotypes 3, 6A/C, 7F, 8, 9N, 10A, 11A, 12F, 15A, 15C, 16F, 17F, 19A, 20A, 22F, 23A, 23B, 24F, 31, 33F, 35B.

[c] Estimated as annual no. nonsusceptible pneumococcal outpatient visits for each nonsusceptibility category divided by annual no. total pneumococcal-attributable outpatient visits (N = 9,722,028 [95% CI 5,969,625, 13,883,036]).

**Table S11. Sensitivity analysis: Burdens of antibiotic nonsusceptible pneumococcal sinusitis assuming serotype distribution based on meta-analysis of studies of pediatric AOM and pneumonia**[8]

| | | Outpatient visits | |
|---|---|---|---|
| **Nonsusceptible to**[a] | **Percent of cases (95% CI)**[b] | *Incidence per 10,000 person-years (95% CI)* | *Annual no. in thousands (95% CI)* |
| First-line antibiotic agent[c] | 10.5 (9.4, 12.0) | 40.1 (24.3, 58.6) | 1,023.9 (621.1, 1,496.9) |
| ≥1 alternative antibiotic agent[d] | 13.9 (12.5, 15.6) | 53.1 (32.2, 77.3) | 1,354.2 (823.1, 1,973.0) |
| >1 antibiotic class | 33.1 (30.9, 35.6) | 126.2 (77.2, 181.7) | 3,221.0 (1,970.0, 4,636.5) |
| ≥3 antibiotic classes | 27.3 (25.3, 29.6) | 104.1 (63.6, 150.1) | 2,656.7 (1,623.2, 3,830.9) |
| Macrolides | 41.4 (38.9, 43.9) | 157.5 (96.4, 226.2) | 4,019.9 (2,460.5, 5,773.6) |
| Antifolates | 25.7 (24.1, 27.4) | 98.0 (59.9, 140.8) | 2,500.2 (1,529.9, 3,593.7) |
| Cephalosporins | 15.4 (14.1, 17.0) | 58.7 (35.8, 84.9) | 1,497.5 (913.0, 2,167.9) |
| Tetracyclines | 19.3 (17.4, 21.4) | 73.4 (44.7, 406.7) | 1,873.7 (1,140.0, 2,723.4) |
| Carbepenems | 12.8 (11.7, 14.1) | 48.7 (29.7, 70.6) | 1,243.8 (757.7, 1,802.6) |
| Penicillins | 15.0 (13.6, 16.8) | 57.3 (34.8, 83.3) | 1,461.9 (889.3, 2,125.1) |
| Lincosamides | 14.0 (12.5, 15.6) | 53.1 (32.3, 77.4) | 1,355.7 (823.6, 1,975.0) |
| Fluoroquinolones | 0.1 (0.1, 0.3) | 0.6 (0.2, 1.3) | 14.1 (5.5, 32.3) |
| Linezolids[e] | 0.0 (0.0, 0.0) | 0.0 (0.0, 0.0) | 0 (0, 0) |
| Glycopeptides[e] | 0.0 (0.0, 0.0) | 0.0 (0.0, 0.0) | 0 (0, 0) |

Abbreviations: CI – confidence interval; No. – number

[a] Nonsusceptibility to antibiotic classes evaluated based on nonsusceptibility to specific agents within each class, as defined in **Table S2**. Linezolids and glycopeptides not shown as all Active Bacterial Core surveillance isolates were susceptible to those classes.

[b] Estimated as annual no. nonsusceptible pneumococcal outpatient visits for each nonsusceptibility category divided by annual no. total pneumococcal-attributable outpatient visits (N = 9,722,028 [95% CI 5,969,625, 13,883,036]).

[c] First-line antibiotic agent for adult sinusitis defined as amoxicillin as proxy for amoxicillin-clavulanate.[6,7]

[d] Alternative antibiotic agents for adult sinusitis defined as doxycycline, levofloxacin, clindamycin, and cephalosporins.[6,7]

**Table S12. Sensitivity analysis burdens of antibiotic-nonsusceptible pneumococcal sinusitis by PCV-targeted serotypes assuming serotype distribution based on meta-analysis of studies of pediatric AOM and pneumonia[8]**

| | | | Outpatient visits | |
|---|---|---|---|---|
| **Nonsusceptible to** | **PCV-targeted serotypes[a]** | **Percent of cases (95% CI)[b]** | *Incidence per 10,000 person-years (95% CI)* | *Annual no. in thousands (95% CI)* |
| 1 first-line antibiotic agent | | | | |
| | PCV15 | 2.0 (1.7, 2.4) | 7.8 (4.7, 11.6) | 198.6 (119.2, 294.8) |
| | PCV20 | 2.1 (1.8, 2.5) | 8.0 (4.8, 11.8) | 203.8 (122.5, 302.4) |
| | PCV21 | 8.9 (8.2, 9.6) | 33.8 (20.7, 48.8) | 863.9 (527.6, 1,245.7) |
| | NVT | 1.5 (0.7, 2.8) | 5.5 (2.3, 11.9) | 139.9 (59.4, 302.8) |
| ≥1 alternative antibiotic agent | | | | |
| | PCV15 | 3.0 (2.5, 3.4) | 11.3 (6.8, 16.6) | 287.3 (173.3, 423.3) |
| | PCV20 | 3.6 (3.1, 4.1) | 13.5 (8.2, 19.9) | 345.8 (209.0, 507.6) |
| | PCV21 | 5.0 (4.5, 5.6) | 19.1 (11.6, 27.7) | 486.7 (296.0, 708.0) |
| | NVT | 8.2 (6.8, 9.9) | 31.1 (18.5, 46.9) | 793.2 (472.0, 1,197.5) |
| >1 antibiotic class | | | | |
| | PCV15 | 4.2 (3.7, 4.7) | 15.9 (9.7, 23.3) | 406.5 (246.6, 593.8) |
| | PCV20 | 5.7 (5.1, 6.4) | 21.8 (13.3, 31.7) | 556.4 (338.4, 808.8) |
| | PCV21 | 16.2 (15.2, 17.5) | 61.9 (37.9, 89.1) | 1,580.2 (966.6, 2,274.0) |
| | NVT | 15.2 (13.3, 17.4) | 57.9 (35.0, 85.0) | 1,478.7 (894.5, 2,170.8) |
| ≥3 antibiotic classes | | | | |
| | PCV15 | 3.5 (3.1, 4.0) | 13.4 (8.1, 19.7) | 343.0 (207.9, 501.8) |
| | PCV20 | 4.6 (4.1, 5.2) | 17.5 (10.6, 25.5) | 447.1 (271.6, 651.2) |
| | PCV21 | 12.7 (11.9, 13.6) | 48.5 (29.7, 69.7) | 1,237.0 (756.8, 1,778.0) |
| | NVT | 13.2 (11.3, 15.4) | 50.3 (30.2, 74.4) | 1,282.7 (772.1, 1,897.9) |

Abbreviations: PCV – pneumococcal conjugate vaccine; CI – confidence interval; No. – number.

[a] PCV15-targeted serotypes: 1, 3, 4, 5, 6A/C, 6B, 7F, 9V, 14, 18C, 19F, 19A, 22F, 23F, 33F. PCV20-targeted serotypes 1, 3, 4, 5, 6A/C, 6B, 7F, 8, 9V, 10A, 11A, 12F, 14, 15B, 18C, 19F, 19A, 22F, 23F, 33F. PCV21-targeted serotypes 3, 6A/C, 7F, 8, 9N, 10A, 11A, 12F, 15A, 15C, 16F, 17F, 19A, 20A, 22F, 23A, 23B, 24F, 31, 33F, 35B.

[b] Estimated as annual no. nonsusceptible pneumococcal outpatient visits for each nonsusceptibility category divided by annual no. total pneumococcal-attributable outpatient visits (N = 9,722,028 [95% CI 5,969,625, 13,883,036]).

[c] First-line antibiotic agent for adult sinusitis defined as amoxicillin.[6,7]

[d] Alternative antibiotic agents for adult sinusitis defined as doxycycline, levofloxacin, clindamycin, and cephalosporins.[6,7]

### SUPPLEMENTAL MATERIAL REFERENCES
